# Long-term Association between NO_2_ and Human Mobility: A Two-year Spatiotemporal Study during the COVID-19 Pandemic in Southeast Asia

**DOI:** 10.1101/2022.10.29.22281700

**Authors:** Zhaoyin Liu, Yangyang Li, Andrea Law, Jia Yu Karen Tan, Wee Han Chua, Yihan Zhu, Chen-Chieh Feng, Wei Luo

## Abstract

Since the COVID-19 pandemic, governments have implemented lockdowns and movement restrictions to contain the disease outbreak. Previous studies have reported a significant positive correlation between NO_2_ and mobility level during the lockdowns in early 2020. Though NO_2_ level and mobility exhibited similar spatial distribution, our initial exploration indicated that the decreased mobility level did not always result in concurrent decreasing NO_2_ level during a two-year time period in Southeast Asia with human movement data at a very high spatial resolution (i.e., Facebook origin-destination data). It indicated that factors other than mobility level contributed to NO_2_ level decline. Our subsequent analysis used a trained Multi-Layer Perceptron model to assess mobility and other contributing factors (e.g., travel modes, temperature, wind speed) and predicted future NO_2_ levels in Southeast Asia. The model results suggest that, while as expected mobility has a strong impact on NO_2_ level, a more accurate prediction requires considering different travel modes (i.e., driving and walking). Mobility shows two-sided impacts on NO_2_ level: mobility above the average level has a high impact on NO_2_, whereas mobility at a relatively low level shows negligible impact. The results also suggest that spatio-temporal heterogeneity and temperature also have impacts on NO_2_ and they should be incorporated to facilitate a more comprehensive understanding of the association between NO_2_ and mobility in the future study.

## 1 Introduction

With over 300 million confirmed cases and 5 million deaths, COVID-19 has spread all over the world since its inception (World Health Organization, 2022). To contain the COVID-19 outbreak, countries had imposed stringent lockdowns and restriction measures since 2020 (Lai *et al*., 2020; R. Zhu *et al*., 2022). Such public health control measures have caused an unintended positive consequence in terms of air quality improvement (Addas and Maghrabi, 2021; Faridi *et al*., 2021).

Sharp declines in air pollution level have been observed during the lockdowns in early 2020 (Kanniah *et al*., 2020; Kumar *et al*., 2020; Tobías *et al*., 2020; Zangari *et al*., 2020; Addas and Maghrabi, 2021; Ghahremanloo *et al*., 2021; Latif *et al*., 2021; Calafiore *et al*., 2022). Faridi *et al*. (2021) reported that the extent of air pollutant reduction may vary with the degree of lockdown measures. Wyche *et al*. (2021) inferred that the NO_2_ decline during the UK lockdown period was due to the 70% vehicle traffic reduction. To investigate the correlation between the observed concurrent declines in NO_2_ and mobility trend, some studies took advantages of the mobility trend datasets (e.g., released by Google and Apple) in interpreting mobility level and demonstrated its significant positive correlation with NO_2_ (Bao and Zhang, 2020; Li and Tartarini, 2020; Zhu *et al*., 2020).

Although this large number of studies have observed NO_2_ level decline and attributed the phenomenon to the reduction in mobility level, those studies focus more on the lockdowns in early 2020. Their investigations that were confined to a short time period for a few months did not account for other neglected factors which could also contribute to the observed pattern. Hence, the observed correlation between NO_2_ and mobility was only limited to this short period and may not persist over a longer time period. Roberts-Semple *et al*., (2021) reported that NO_2_ exhibits seasonal characteristics in northeastern New Jersey. Zangari *et al*. (2020) found that the NO_2_ level in New York City decreased between January and May in each year since 2015, with or without the lockdown in early 2020. In addition to temporal changes, mobility and NO_2_ levels vary across regions and countries. By adopting the entire 2020 data in Singapore, Li *et al*. (2022) did confirm that it would not only overestimate the correlations between NO_2_ and mobility if only focusing on the lockdown time period, but show significant spatial variations in the correlation between NO_2_ and mobility as well. Thus, there is a demanding need to understand the spatio-temporal heterogeneity of NO_2_ and mobility over a larger geographical area and a longer time period.

NO_2_ is known to cause problems on environment and health (Tian *et al*., 2019). Despite various mitigation measures implemented, such air pollutant has been a long-standing issue in Southeast Asia (SEA) (Kanniah *et al*., 2020). During the COVID-19 pandemic, countries in SEA had imposed stringent lockdowns in 2020 along with various restriction measures (e.g., remote working, curfew, restrictions on social gathering and traveling) which limited human movement and activities to a great extent (Luo *et al*., 2022). These changes in human behaviours provided an unprecedented opportunity to study how decrease in citizens’ mobility affects air pollution (Piccoli *et al*., 2020). Thus, this study aims to perform spatio-temporal analyses of NO_2_ level and mobility in SEA from March 2020 to February 2022 and develop a NO_2_ prediction model to explore its response to key influencing factors. Different from previous studies, we utilized high-resolution mobility and NO_2_ data in a two-year time period to investigate the long-term NO_2_ response in SEA. The study aims to 1) identify the temporal changes of NO_2_ and mobility throughout the two years of COVID-19 pandemic (March 2020 – February 2022); 2) evaluate the correlations between NO_2_ and mobility; and 3) assess the impact of different factors on NO_2_.

## 2 Materials and methods

### 2.1 Study area

Our study area consisted of 10 countries within SEA: Brunei, Cambodia, Indonesia, Laos, Malaysia, Myanmar, the Philippines, Singapore, Thailand, and Vietnam. We extracted and analysed NO_2_ level, mobility, and other factors including travel modes, temperature, and haze from March 2020 to February 2022. This period was chosen because of reduced mobility due to government-mandated COVID-19 measures that occurred in different intensities and timescales across the countries, enabling direct comparison of mobility impact on NO_2_ levels. The whole study area of 770 km^2^ was square gridded to 0.25º for the follow-up analyses (Figure 1).

**Figure 1.**
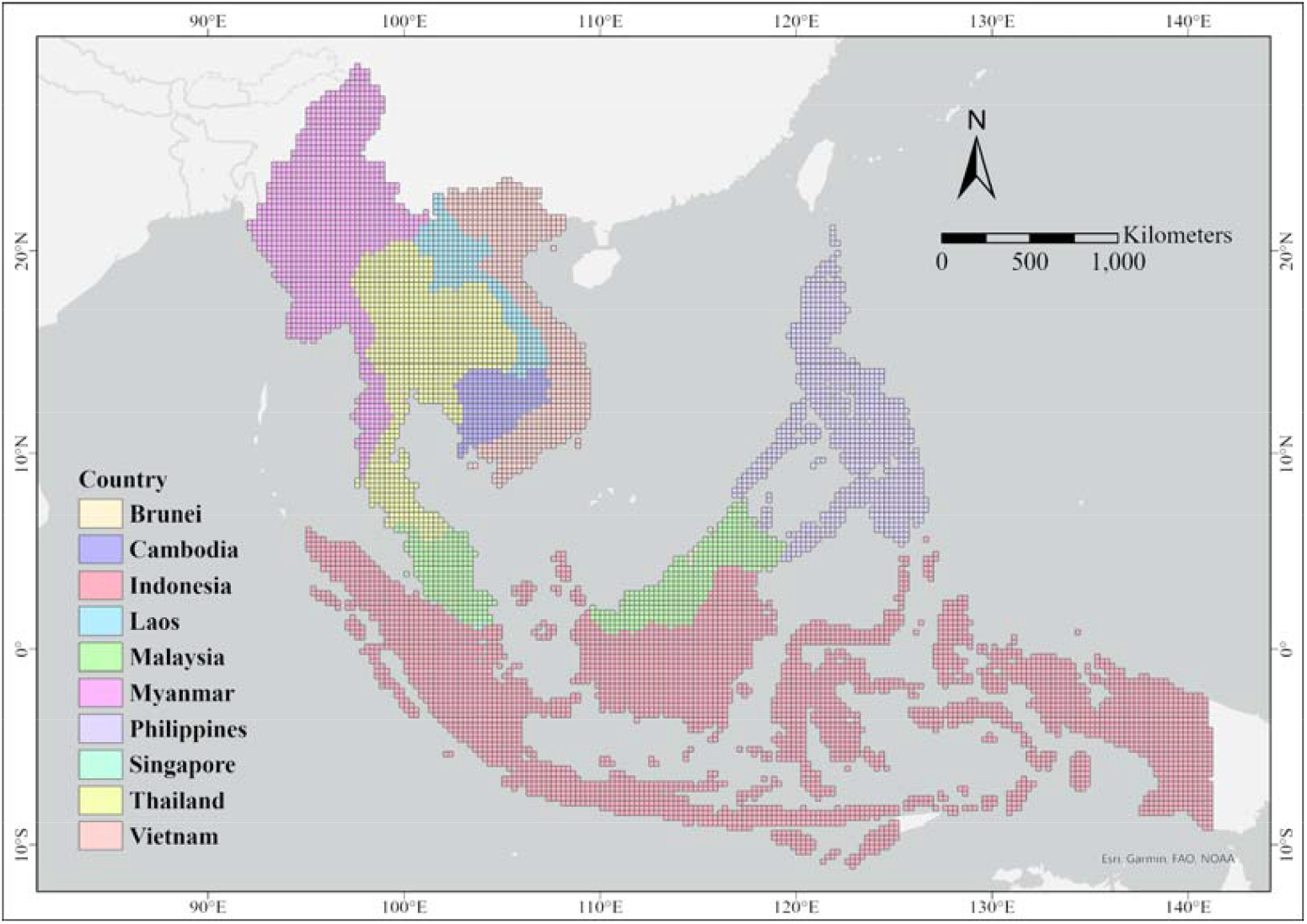
Area of study with coloured grids (0.25º × 0.25º) representing the different countries of interest within SEA.

### 2.2 Data sources and pre-processing

#### 2.2.1 NO_*2*_

The level-3 product of daily NO_2_ observations (Tropospheric NO_2_ column density) of TROPOMI/Sentinel-5 was obtained from the Google Earth Engine cloud-based platform. The original data was resampled from the source resolution of 0.01º to 0.25º through mean operator.

#### 2.2.2 Mobility

This study adopted mobility data from Facebook Data for Good at Meta (Maas, 2019) and Apple Mobility Trends Reports (Apple, 2020). The Facebook movement dataset records user movements between origin and destination tiles aggregated using Facebook users’ locations every eight hours (8:00, 16:00, 00:00). As the resolution of Facebook’s data in different countries varied from level 11 to level 14 (approximately 0.09° to 0.17°) in Bing tile system (Schwartz, 2018). We aggregated the original dataset to 0.25° grids by aggregating the mobility volumes within each grid (data records with 0m distance were excluded). As the dataset only contains information of the origin and destination rather than the actual movement route, it is difficult to assign movement value to other grids apart from the origin and destination grids, especially in long-distance travels. To minimize the inherent uncertainties in longer distance movements, the subsequent analyses are limited to movements whose origin and destination belong to the same or neighbouring grids (accounting for 93.33 percent of the total movement volume). The corresponding Facebook movement values were assigned equally to both starting and ending grids. Apple moving trends present the percentage changes of two travel modes, driving and walking over time. Note that, Apple moving trends data is calculated by referring to the number of searches carried out on its Maps application.

Meteorological conditions were reported to affect pollutant concentration (Pearce et al., 2011; He et al., 2016; Wang et al., 2017). We sourced six meteorological parameters that could potentially influence NO_2_ level (Jiang et al., 2014; He et al., 2017) from 1st March 2020 to 31st December 2021: 1) maximum hourly 2m surface temperature, 2) dew point temperature, 3) mean hourly 10m u-component of wind, 4) 10m v-component of wind, 5) hourly precipitation, and 6) surface pressure. More specifically, we extracted each of these daily parameters from the ERA5 hourly reanalysis dataset from the Copernicus Climate Data Store (Copernicus, 2022) and aggregated them into daily meteorological values respectively.

#### 2.2.3 Haze

Google Trends, an open-source database, was used to retrieve the indexed popularity of Google searches indicating the public’s interest on that topic (Google, 2022). The English word ‘haze’ was used to understand the severity of haze at country level. We utilised “pytrends” (an Application Programming Interface for Google Trends) to extract and scale datasets obtained for different countries, providing compatible datasets in both space and time (General Mills, 2022). While the values are indexed, which limits our ability to determine the actual number of searches, the set of varying values remains a useful proxy for the severity of haze in that location. Other keywords within and across the national languages were used in the search, but the results were noisy and excluded. For instance, searches for ‘transboundary haze’ were only detected in Singapore, Malaysia, and Indonesia, possibly denoting its prevalence only in those countries, or an issue of lexicons instead. Hence, only the English word ‘haze’ was employed as an approximation to the presence and severity of haze.

### 2.3 Data analyses

To address our study objectives, we incorporated three synergistic analyses: (1) a case study of Indonesia; (2) Space-Time Cube (STC) and Emerging Hotspot Analysis (EHSA) in SEA; (3) a Multi-Layer Perceptron (MLP) model to predict NO_2_ and assess the impact of different input factors on NO_2_. Due to the constraint of available data from Facebook movement dataset, we conducted the first two analyses with limited settings (Table 1). For the first analysis, we conducted a case study spanning a long time period to identify the temporal changes throughout the COVID-19 pandemic. Nevertheless, the complete dataset of 2020 is available only in Indonesia and Singapore. We chose to carry out our case study in Indonesia considering that it has a much larger spatial area than Singapore. The analysis expands to 10 countries in SEA from May 2021 because they are available for the whole of SEA. For the third analysis, our study covers a period between 1st April 2020 and 31st December 2021 because the meteorological data of 2022 was unavailable at the time this study was conducted.

**Table 1.**
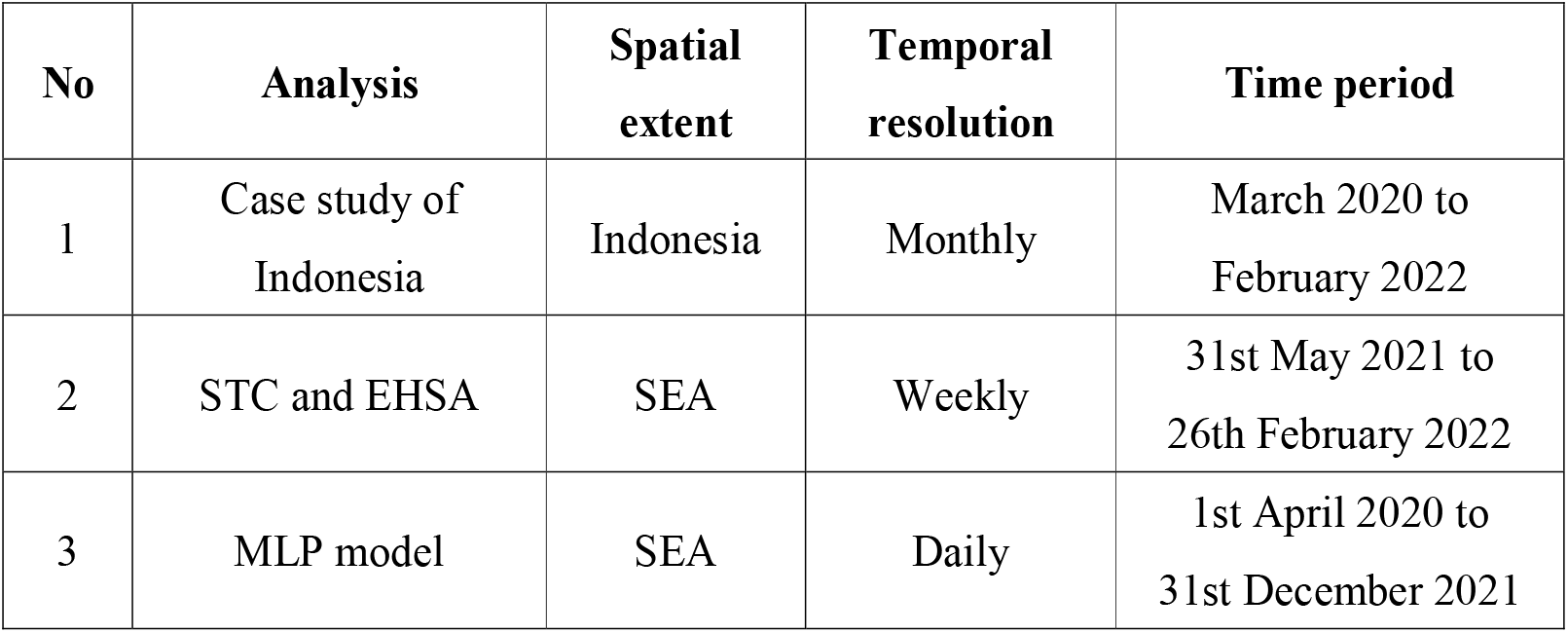
An overview of the corresponding spatial extent, temporal resolution and time period for each analysis.

#### 2.3.1 Indonesia case study

As the most populous country in SEA, the case study of Indonesia was conducted to provide an initial and exploratory understanding of the linkage of mobility and NO_2_ variations between March 2020 and February 2022. The monthly NO_2_ data and Facebook movement data in a spatial resolution of 0.25° were studied across Indonesia. Ordinary kriging (Wackernagel, 2003) was applied to fill in the grids without data (Childs, 2004; Shad *et al*., 2009). A series of continuous raster maps were generated to facilitate the identification and subsequent interpretation of potential patterns in Indonesia.

#### 2.3.2 Space-time hotspot analysis in SEA

We used the EHSA tool in ArcGIS Pro 2.8 software (Esri, 2022) to detect spatio-temporal trends in NO_2_ and Facebook movement within their respective STC. We conducted this analysis at a weekly interval between 31st May 2021 and 26th February 2022 over the 10 countries in SEA. We then examined spatial clusters by hot spot categories in greater detail through examining Pearson’s correlation between the log of NO_2_ level and the log of Facebook movement values over the 39-week period for each point in a spatial cluster. As the magnitude of the ranges of NO_2_ (-5.10e-5 to 8.26e-4) and Facebook movement (2.50 to 8.28e+7) could potentially skew the analysis outcome, we performed data cleaning and transformation as below. Firstly, NO_2_ data points with less than 1e-6 mol/m^2^ and Facebook movement values below 50 were excluded as they were observed as noise (Figure 2). Then we conducted a logarithmic transformation of base 10 for both variables. Note that before taken logarithm, the filtered NO_2_ data were multiplied by 10e+7 to ensure that data values remain positive. By removing data anomalies and reducing data skewness, these transformations improve the accuracy of our analyses.

**Figure 2.**
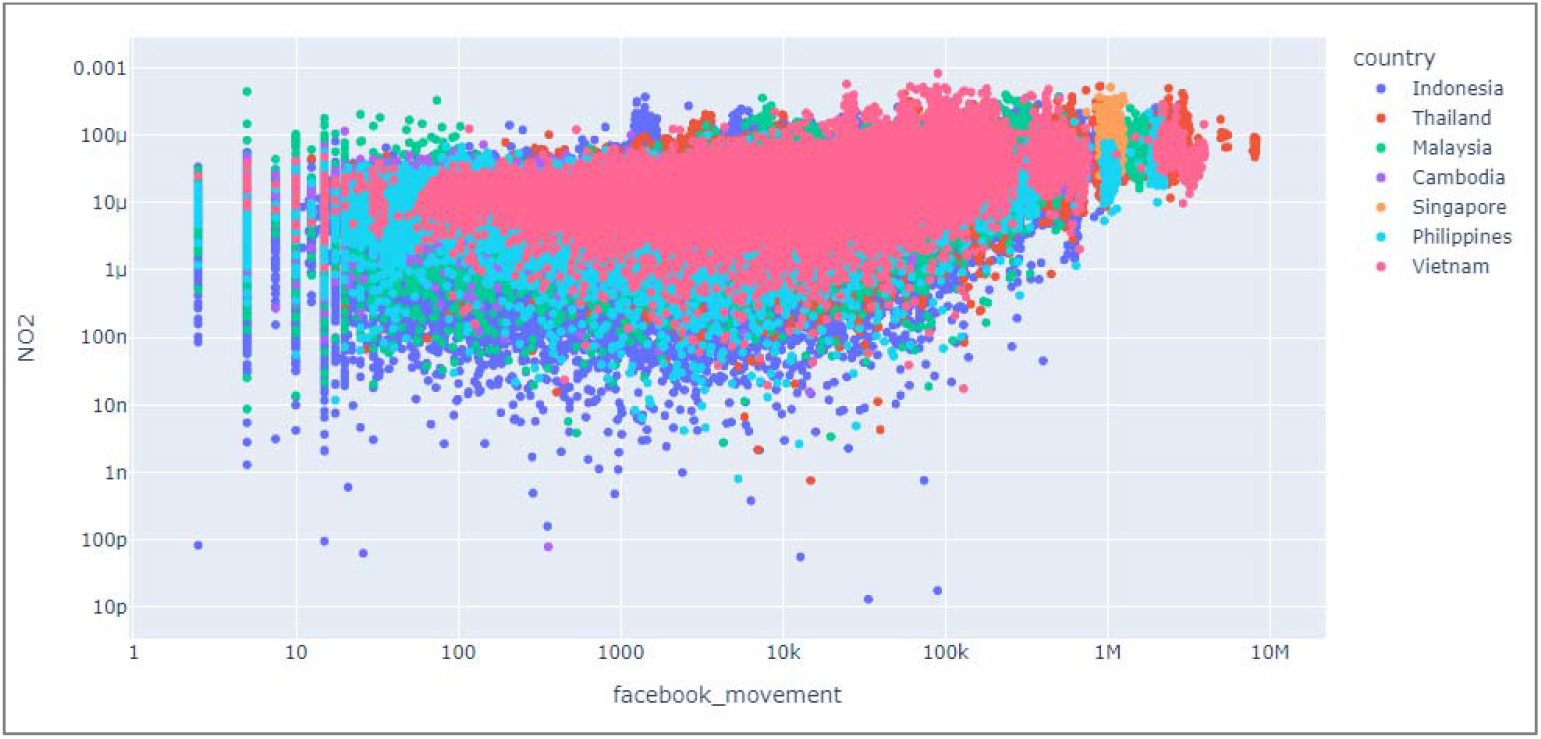
Scatter plot between NO_2_ and Facebook movement.

#### 2.3.3 NO_2_ prediction in SEA using MLP model

In addition to the correlation between NO_2_ and mobility, we evaluated the impacts of different factors on NO_2_ using an MLP model. MLP is the most popular feedforward model with good performance in air pollution prediction and forecasting (Cabaneros *et al*., 2019; Shams *et al*., 2021). It is an artificial neural network (ANN) that consists of an input layer, an output layer, and multiple hidden layers in between. The number of nodes in the input layer should be the same as the number of input variables. Input variables included 15 parameters (Table 2) across five categories: location (i.e., longitude and latitude), elapsed days (i.e., the number of days since the start of the study period), meteorological parameters (i.e., rainfall, wind speed, u-component of wind, v-component of wind, temperature, dew-point temperature, surface pressure), haze, and mobility (i.e., Facebook movement, the logarithm of Facebook movement, Apple driving and walking trends). To improve the model performance of MLP, we adopted the same method for data transformation as mentioned in section 2.3.2.

**Table 2.** List of features in MLP model.

| **Feature category** | **Feature meaning** | **Feature name** |
| --- | --- | --- |
| Location | Longitude | lon |
|  | Latitude | lat |
| Elapsed days | The number of days since the start of study period | day |
| Meteorological parameters | rainfall | rainfall |
|  | wind speed | wind speed |
|  | u-component of wind | u-wind |
|  | v-component of wind | v-wind |
|  | temperature | 2m-temp |
|  | dew-point temperature | dew-pt |
|  | surface pressure | surface-p |
| Haze | relative number of haze searches | haze |
| Mobility | Facebook movement | facebook_movement |
|  | logarithm of Facebook movement | log_facebook_movement |
|  | Apple driving trend | apple_driving |
|  | Apple walking trend | apple_walking |

We used the rectified linear unit (ReLU) activation function in this MLP model. ReLU outputs negative values to 0 and keeps other values the same as the inputs. Therefore, we converted all the negative values to positive values using the transformation method as mentioned in 3.1.2. to improve the model efficiency. By training different MLP models, we could then determine the optimum model architecture to be used in this study that resulted in a smaller error with little additional computational effort. Of all the data records, 70% were used for training and while the remainder were used for testing. Each input variable was normalised before training the model. Root mean square error (RMSE), mean absolute error (MAE) and R^2^ were used as the metrics for model evaluation.

We then used SHapley Additive exPlanation (SHAP) to explain the model in order to quantify the impact of the different features on the model output (Lundberg and Lee, 2017; Li, 2022). Traditional sensitivity analyses usually evaluate the changes in the output as a result of the change in only one input parameter. This may not be realistic as the possible correlation between variables is not accounted for. For example, air pressure and air temperature tend to increase or decrease together. SHAP values can account for the possible interactions amongst the input features when evaluating the impact of each feature on the output. However, it should be noted that the SHAP value or impact is different from the correlation. A high correlation means that the two variables tend to change together, while a higher SHAP value indicates a greater impact of one variable on the other, i.e., a change in one variable results in a greater magnitude of change in the other. The SHAP results can pinpoint the parameters that have relatively higher impacts on the NO_2_ level in SEA.

## 3 Results & Discussion

### 3.1 Spatio-temporal distribution of NO_2_ and mobility level in Indonesia

To begin with, we compared the spatio-temporal distribution variations of NO_2_ and Facebook movement levels in Indonesia during March 2020 and February 2022. Overall, the result showed a decreasing trend during the study period except in March 2021 (Figure 3). On the other hand, NO_2_ showed a cyclical trend in 2020 and an increasing trend in 2021 before reaching peak in October 2021.

**Figure 3.**
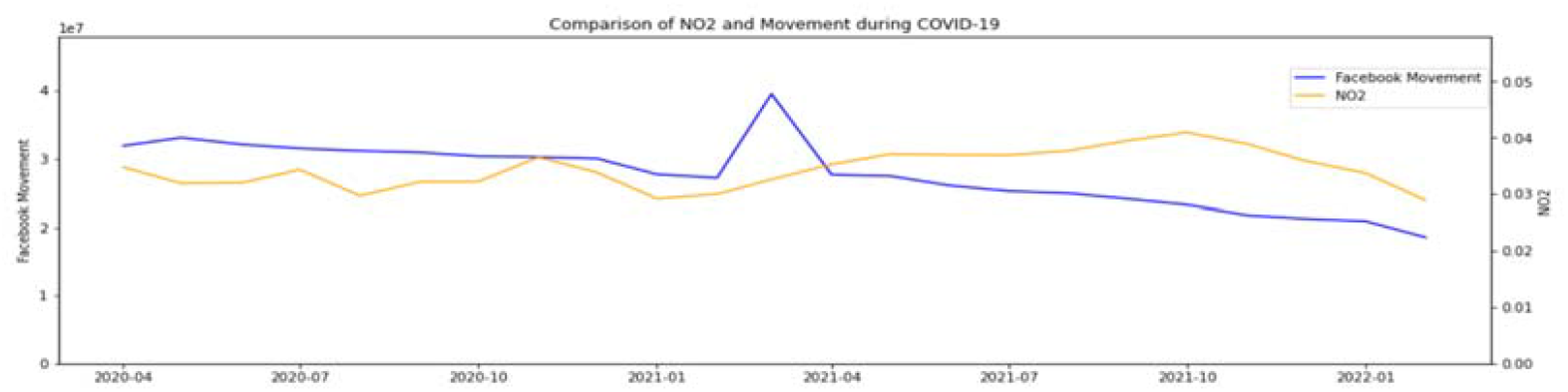
Comparison of Facebook movement and NO_2_ in Indoensia.

South and West Indonesia showed high NO_2_ levels, especially in Jakarta and its surrounding regions where the maximum mean NO_2_ value reaches 1.01e-04 mol/m^2^ in April 2020 (Figure 4). In comparison, the NO_2_ level is substantially lower in other areas, especially in East Indonesia. Facebook movement presents a similar distribution. The similarity in distributions of Facebook movement and NO_2_ level suggests a possible association between them.

**Figure 4.**
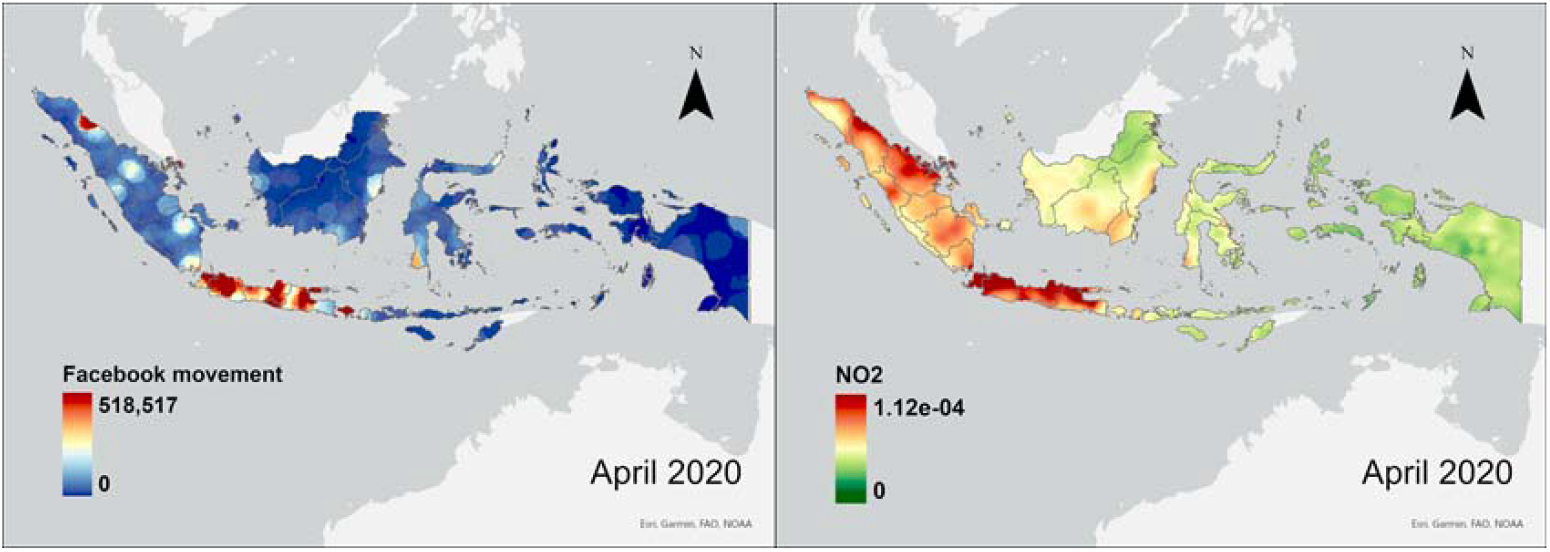
Spatial Distribution of Facebook movement and NO_2_ level in April 2020.

Figures 5 and 6 illustrate the temporal changes in Facebook movement and NO_2_ distribution. Maps showing the temporal variations over the last two years are presented in Appendix A. During the first wave of COVID-19, although Facebook movement did not vary significantly, NO_2_ exhibited an oscillating pattern. Similarly, in 2021, the Facebook movement constantly decreased ever since March whereas the NO_2_ level did not present a decreasing trend. Moreover, we observed an increase in NO_2_ over the months in regions including Bali, Jawa Barat, Lampung and Kalimantan Barat. Overall, the temporal variation of NO_2_ and Facebook movement did not present similarity. This result contradicts previous studies in which the reduction of mobility had led to a decrease in NO_2_ level (Sicard *et al*., 2020; Faridi *et al*., 2021). Therefore, we hypothesise that human movement volume may not be the single significant factor affecting air pollution. For instance, forest fires had occurred in Indonesia causing air pollution and dryness which pre-matured the onset of haze in early 2021 (Hicks, 2021).

**Figure 5.**
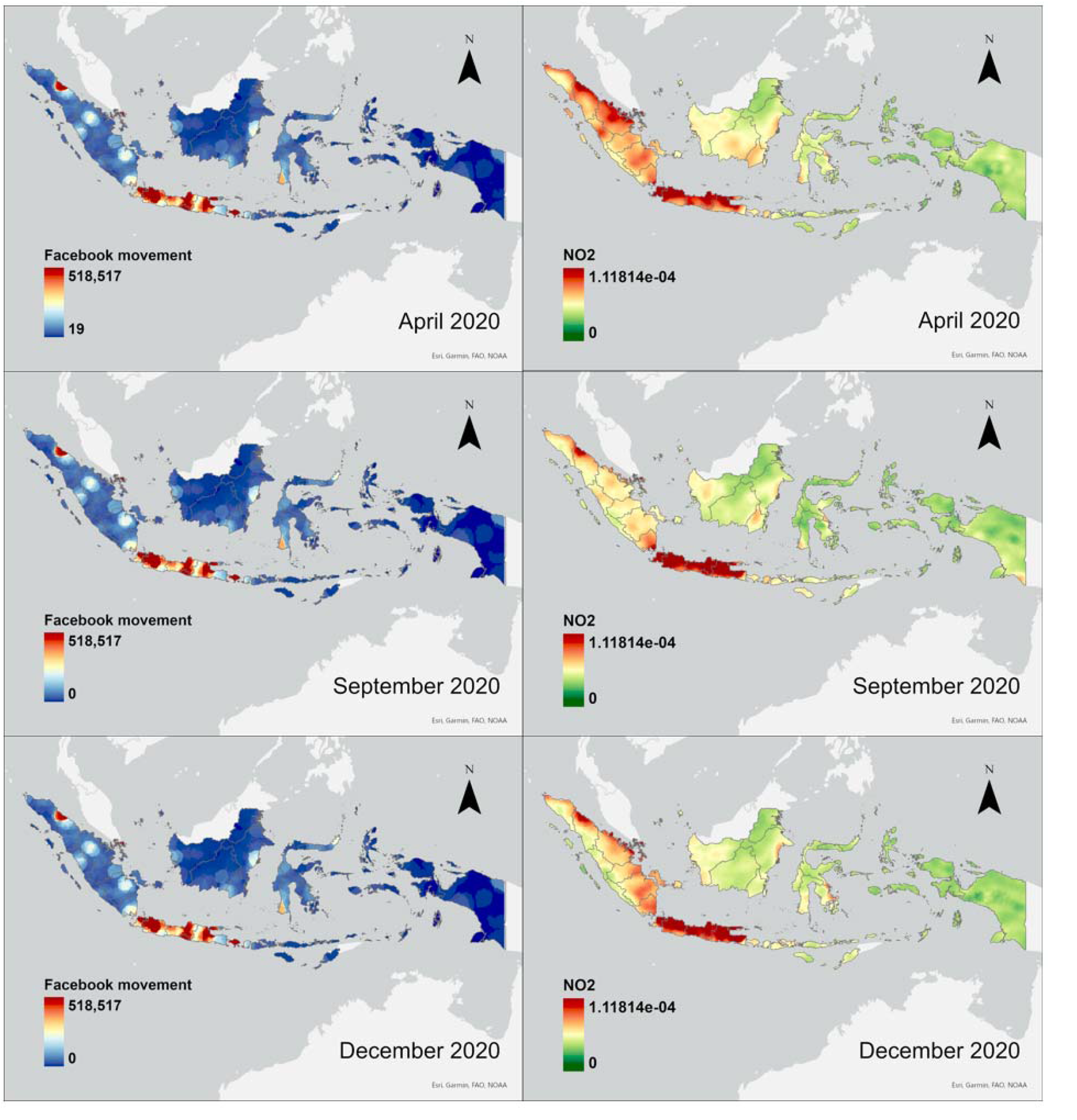
Comparison of Facebook movement and NO_2_ level during the first wave (April 2020 – December 2020) of COVID-19 in Indonesia.

**Figure 6.**
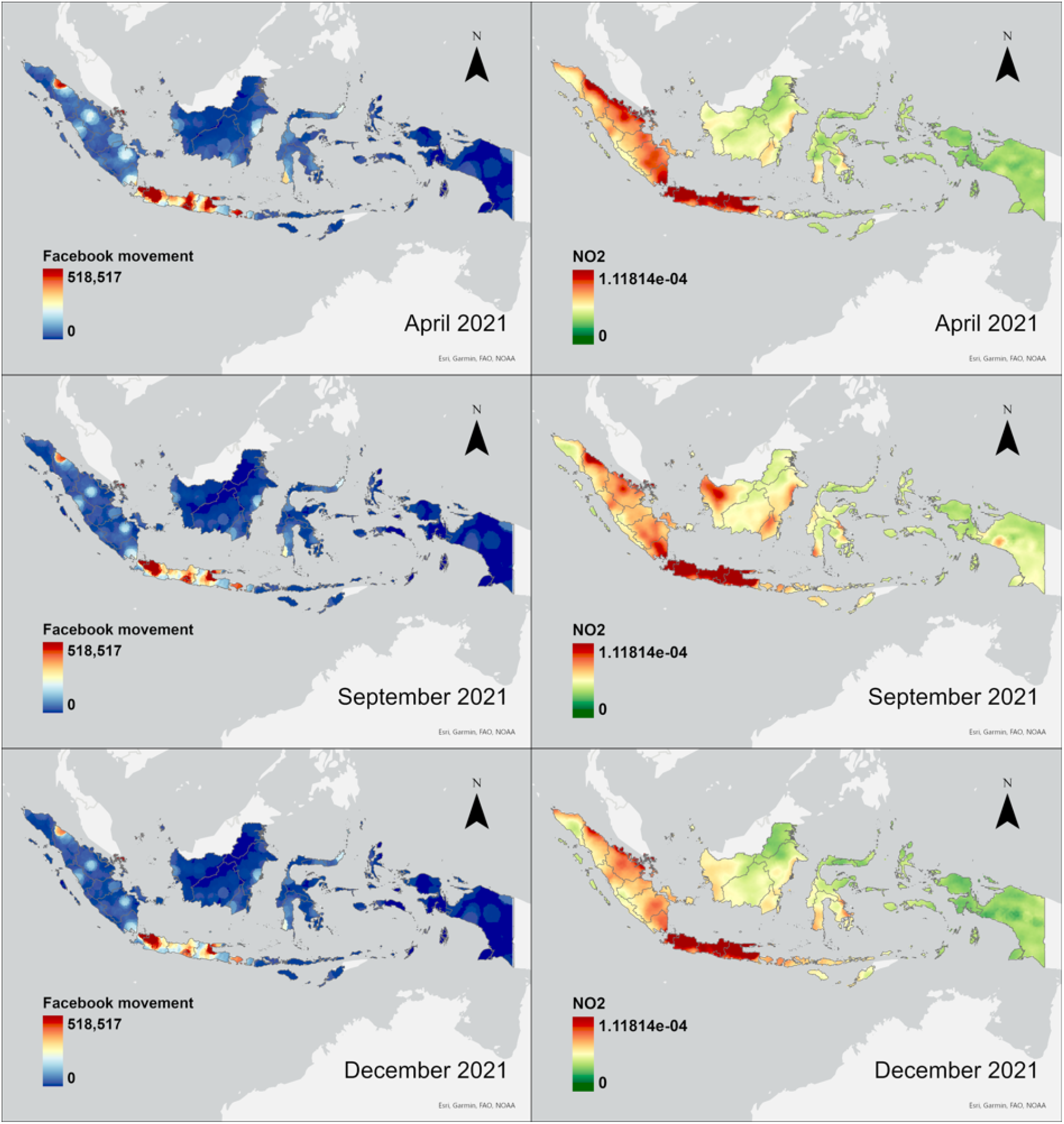
Comparison of Facebook movement and NO_2_ level during the second wave of COVID-19 (April 2021 – December 2021) in Indonesia.

### 3.2 Spatio-temporal evaluation of the association between NO_2_ and Facebook movement trends in SEA

Subsequently, we conducted analysis using EHSA at a finer temporal scale (weekly) in all selected SEA countries. Due to insufficient Facebook movement data, the study period for this analysis was limited to 31st May 2021 and 26th February 2022. Figure 7 shows the temporal changes in Log(NO_2_) and Log(Facebook movement) during the study period. Similar to previous findings, there was a consistent pattern of a slight decrease in Facebook movement throughout the 39-week study period with a sharp decrease approximately at the 25^th^ week in all eight SEA countries (Figure 7). On the other hand, we found no consistent patterns in NO_2_ levels over time. Nonetheless, there was a slight increase in Log(NO_2_) approximately around the 20^th^ week and the 25^th^ week in Thailand and Myanmar respectively.

**Figure 7.**
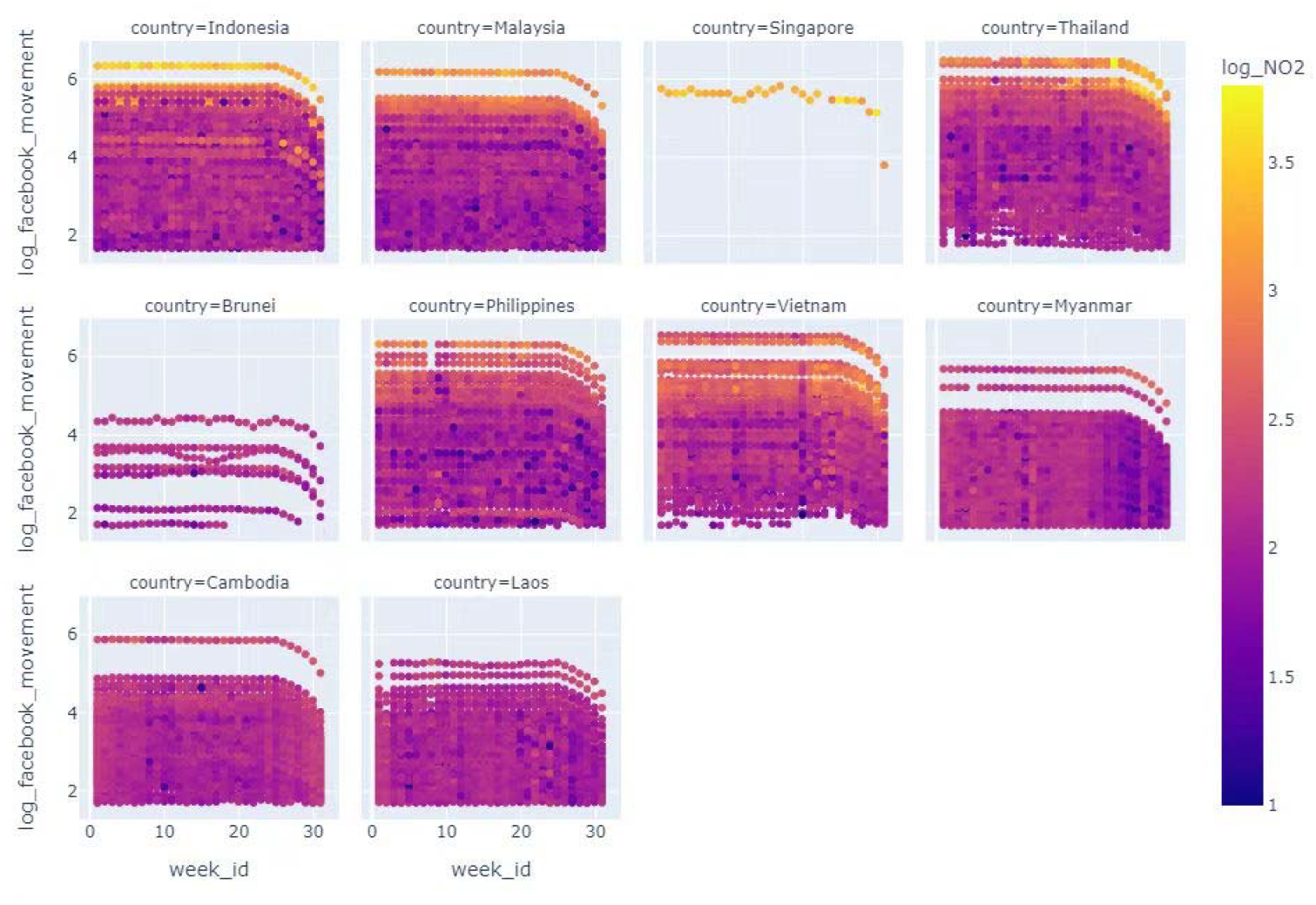
Change in Log(facebook_movement) against Log(NO_2_) between June 2021 and February 2022. Each circle in the figure corresponds to a grid in the country. The darker the circle, the lower the Log(NO_2_).

From the complete EHSA result derived from the NO_2_ level (Figure 8a), it is observed that hot spots were detected mainly in the northwest of SEA. In comparison, central and south of SEA, which were mainly covered by Malaysia and Indonesia, were dominated by cold spots. Several hot spot patterns were identified, including new, intensifying, persistent and diminishing hot spots (Figure 8b). In terms of the spatial-temporal patterns of NO_2_, the diminishing hot spots indicate decreasing clustering intensity of the NO_2_ space-time hotspots in a statistically significant trend over time. Contrastingly, the intensifying hot spots showed a statistically significant increase in clustering intensity over time.

**Figure 8.**
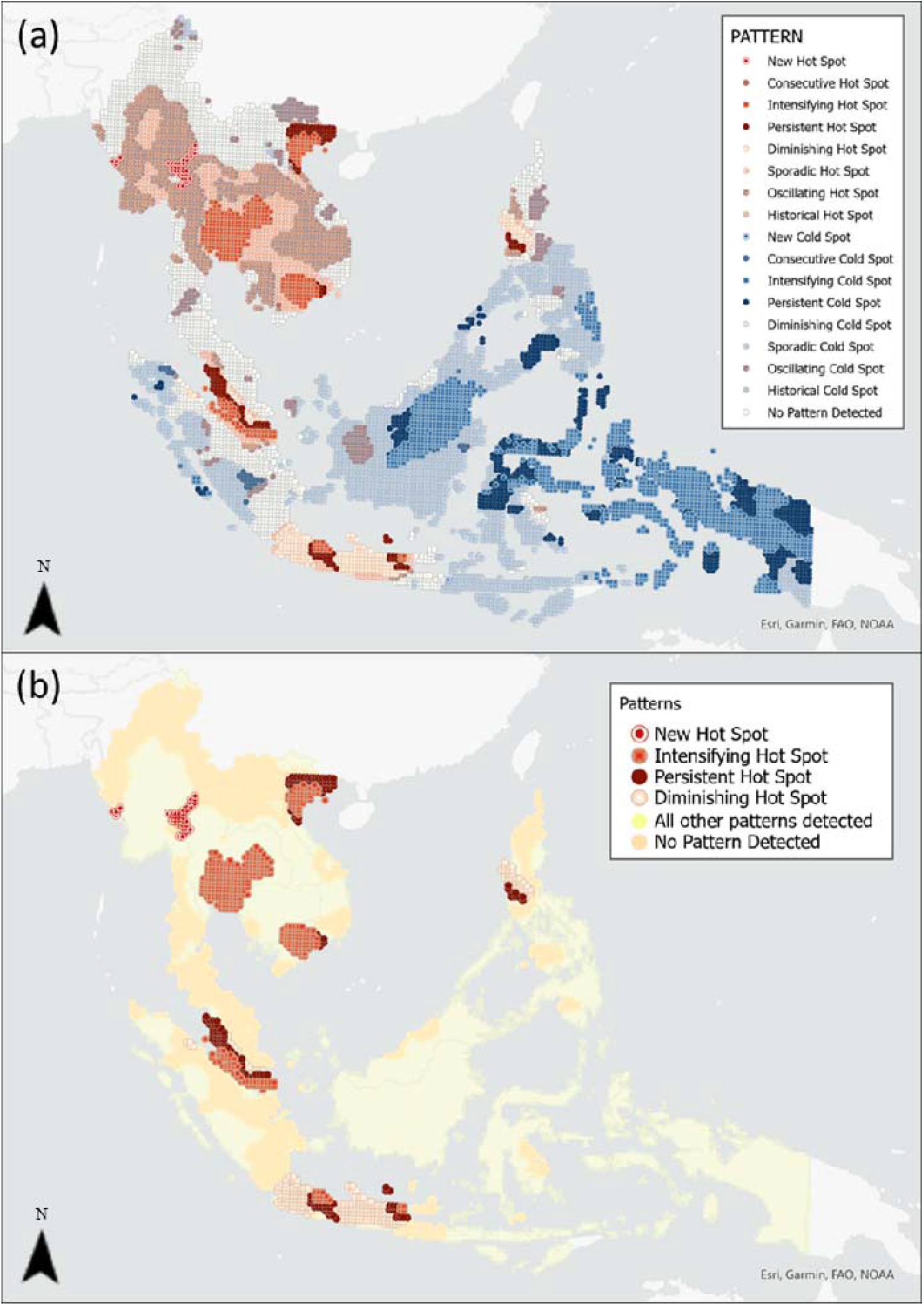
(a) Complete result of NO_2_ EHSA result and (b) result of NO_2_ EHSA with only selected patterns of interest.

The complete result of Facebook movement EHSA (Figure 9) is different from that of NO_2_ EHSA (Figure 8a). The Facebook movement EHSA result showed substantially fewer observed patterns, where only diminishing, sporadic and historical hot spots but no cold spots were detected. In the context of Facebook movement, diminishing hot spots indicate that locations of statistically significant mobility hot spots exhibit decreasing clustering pattern. On the other hand, the sporadic hot spots indicate locations where statistically significant hot spots were only identified at some inconsistent time steps. Lastly, historical hot spots indicate locations where the level of mobility has been detected as statistically significant hot spots for the major time steps in the past. These hot spots are observed mostly in the major SEA cities such as Hanoi, Jakarta, Johor and Singapore, where lockdown restrictions would influence mobility more due to higher population and more intense economic activities (Heroy et al., 2021). This corroborates with the overall decrease in Facebook movement over the entire study period (Figure 7).

**Figure 9.**
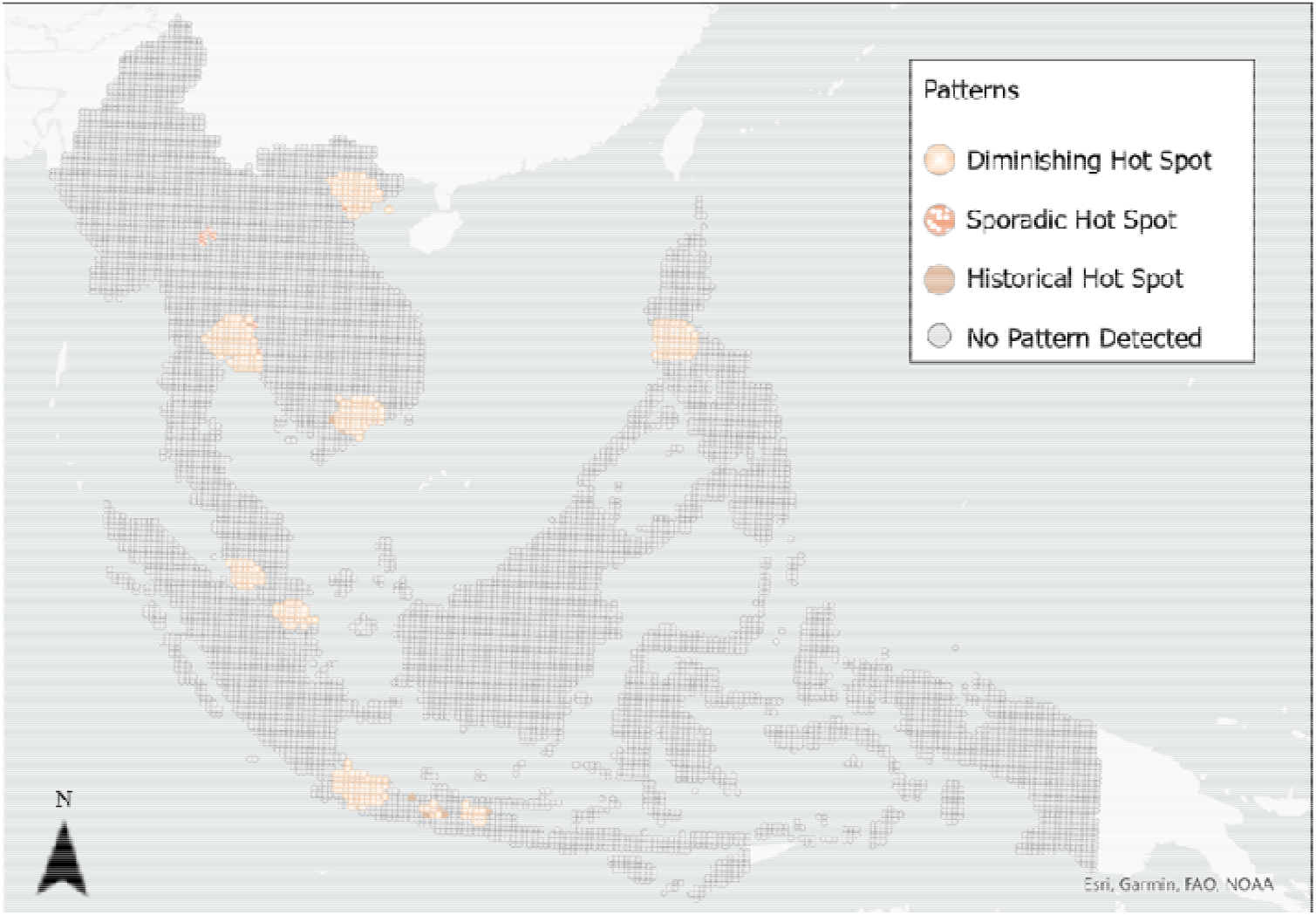
Complete EHSA result of Facebook movement.

As this study seeks to examine the effect of mobility changes on NO_2_, we conducted correlation analyses on specific spatio-temporal patterns with reference to the EHSA results of NO_2_. While several types of NO_2_ spatio-temporal hot spots were identified using EHSA, the correlation between Log(NO_2_) and Log(Facebook movement) varied widely geographically from June 2021 to February 2022. Such variation was observed within a country and even within a city.

In the case of the intensifying hot spots, while positive correlation between Log(NO_2_) and Log(Facebook movement) was observed in Chon Buri province, Thailand, a mix of positive and negative correlations were observed in the Rayong and Chanthaburi provinces, Thailand (Figure 10). This trend did not support the previous findings that the degree of movement restriction correlates positively with NO_2_ level. In fact, the same spatial cluster of NO_2_ intensifying hot spots in Chon Buri and Rayong were identified as diminishing hot spots or no pattern detected in the Facebook movement EHSA result (Figure 9). While NO_2_ levels at these locations became stronger hot spots over time, the corresponding mobility hot spots became less clustered. This contrasting observation further confirms that there is lack of evidence supporting a direct association between mobility and the resultant NO_2_ levels.

**Figure 10.**
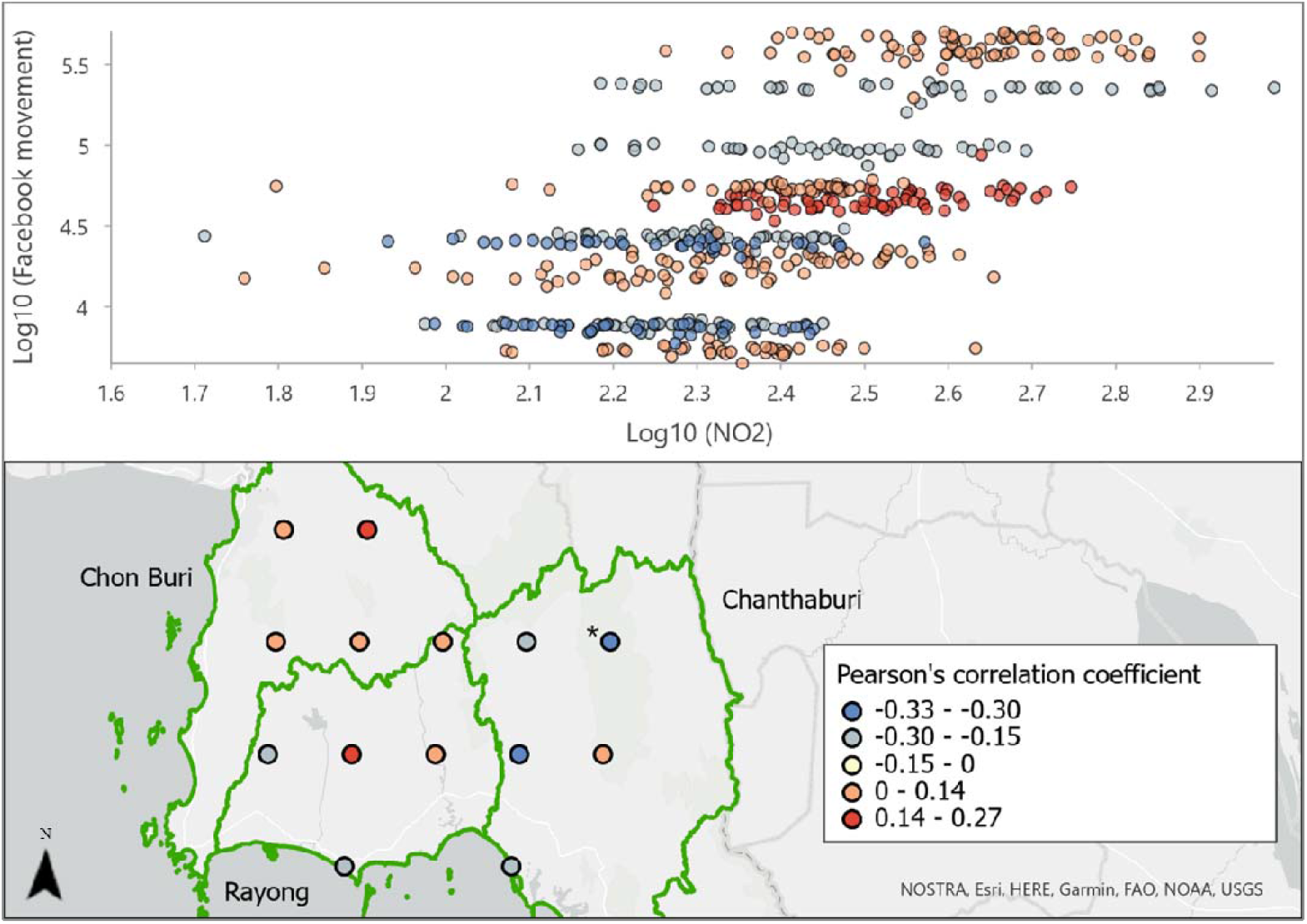
Correlation between Log(Facebook movement) and Log(NO_2_) in intensifying hot spots in Thailand.

Variation in correlation was also observed among the persistent hot spots in the provinces around Ha Noi in Vietnam (Figure 11), and the new hot spots were found along the border between Thailand and Myanmar (Figure 12). While the persistent hot spots indicated statistically significant higher NO_2_ levels relative to their space-time neighbours over most of the 39 weeks without discernible trends, the new hot spots indicate a statistically significant space-time hotspot only in the last time step and not in any of the previous ones. Yet, the weak positive correlation and moderate negative correlations between Log(NO_2_) and Log(Facebook movement) show insubstantial evidence to attribute the changes in NO_2_ levels to the changes in mobility. Likewise, these locations were identified as the diminishing hot spots or absence of space-time pattern in the Facebook movement EHSA result (Figure 9). This similarly suggests that while mobility hot spot intensity decreased in these points, the NO_2_ hot spot intensity stayed relatively constant over time.

**Figure 11.**
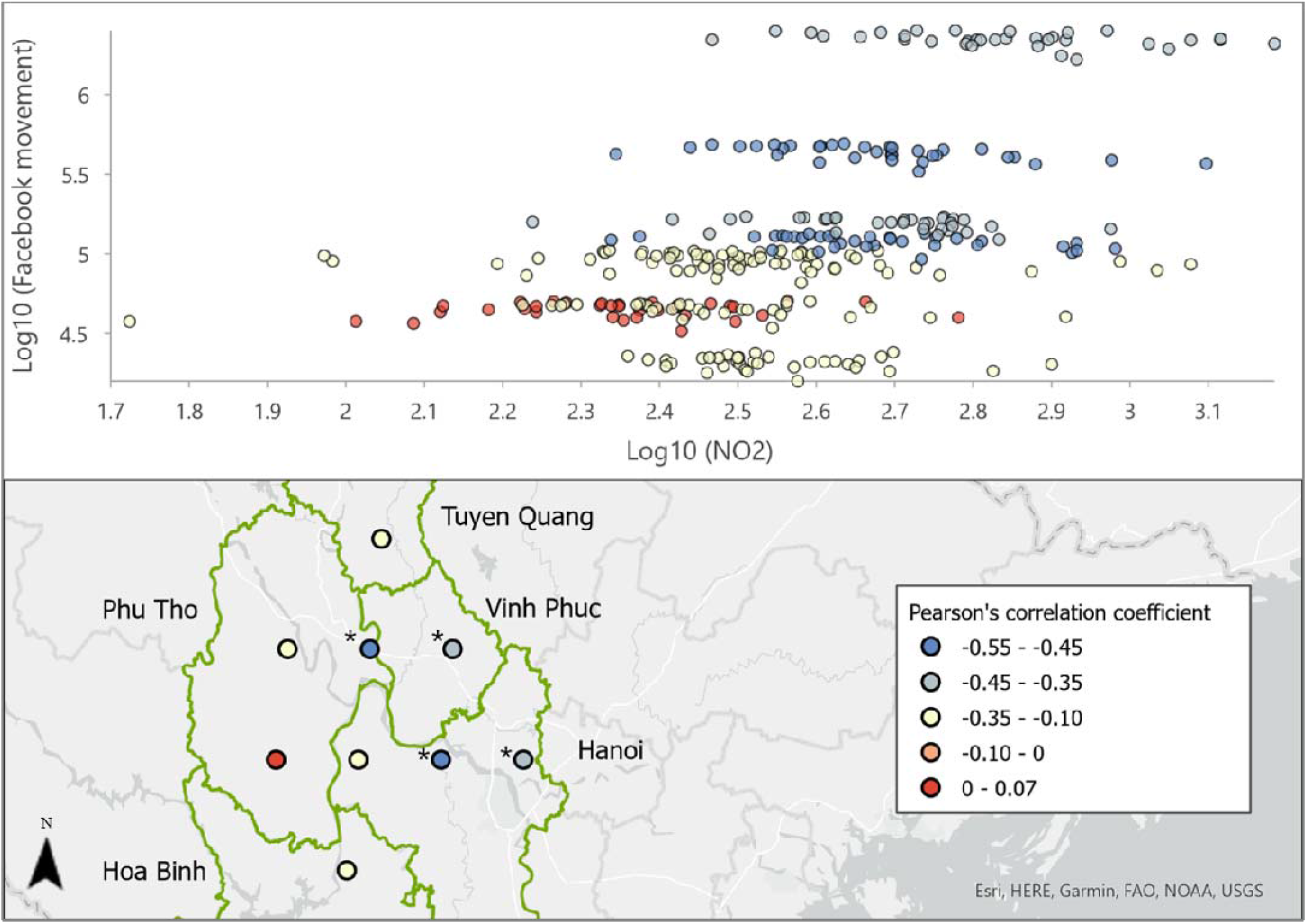
Correlation between Log(Facebook movement) and Log(NO_2_) in persistent hot spots in Vietnam.

**Figure 12.**
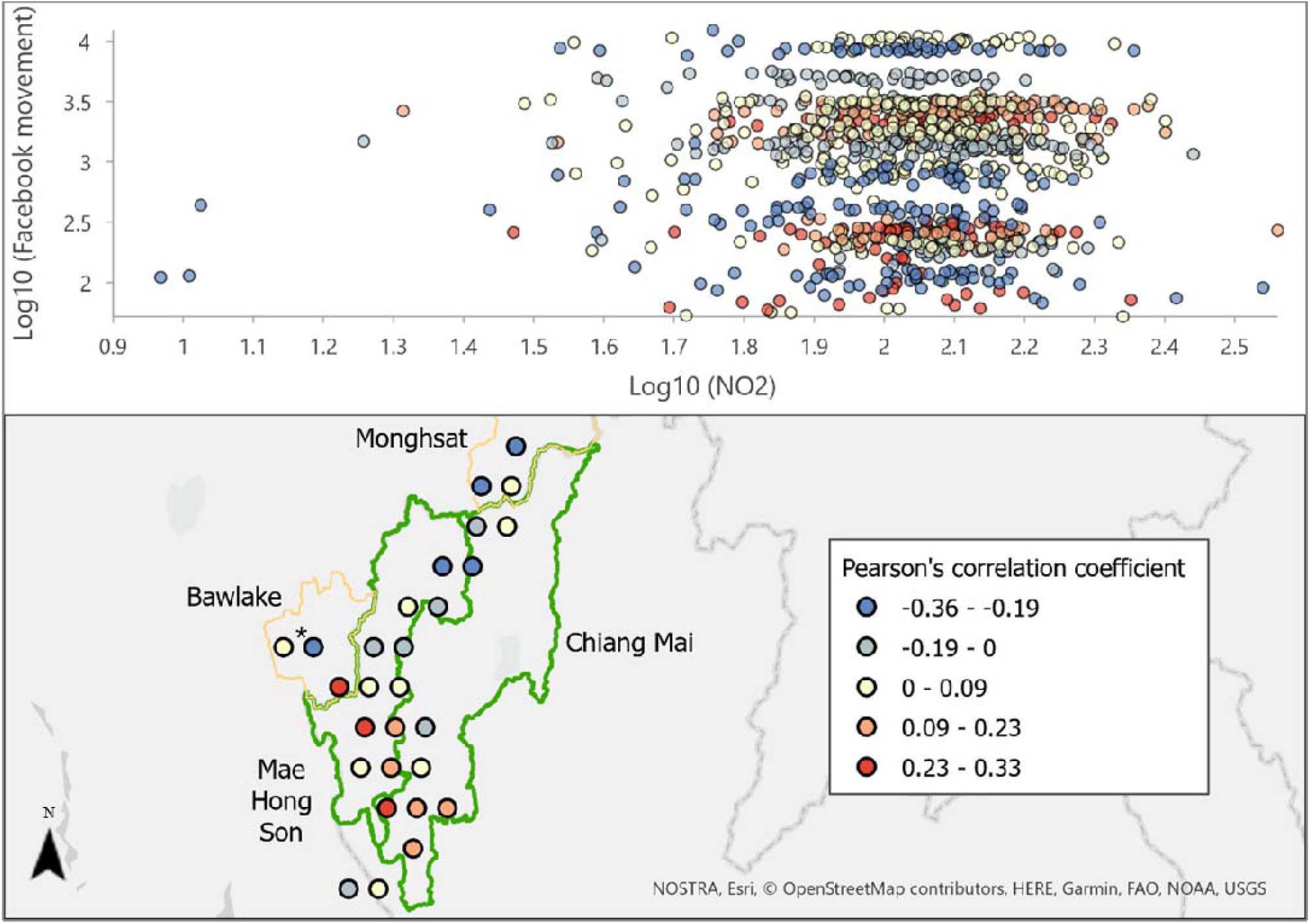
Correlation between log (Facebook movement) and log (NO_2_) in new hot spots in Thailand and Myanmar.

In the case of the diminishing hot spots, we detected two clusters of points in Indonesia (Figure 13) and Malaysia (Figure 14), which exhibit positive correlations between Log(NO_2_) and Log(Facebook movement). In addition, the corresponding points were also identified as the diminishing hot spots in the EHSA results of the Facebook movement. This suggests that both NO_2_ levels and Facebook movement decrease over time, validating the positive correlation between these two variables.

**Figure 13.**
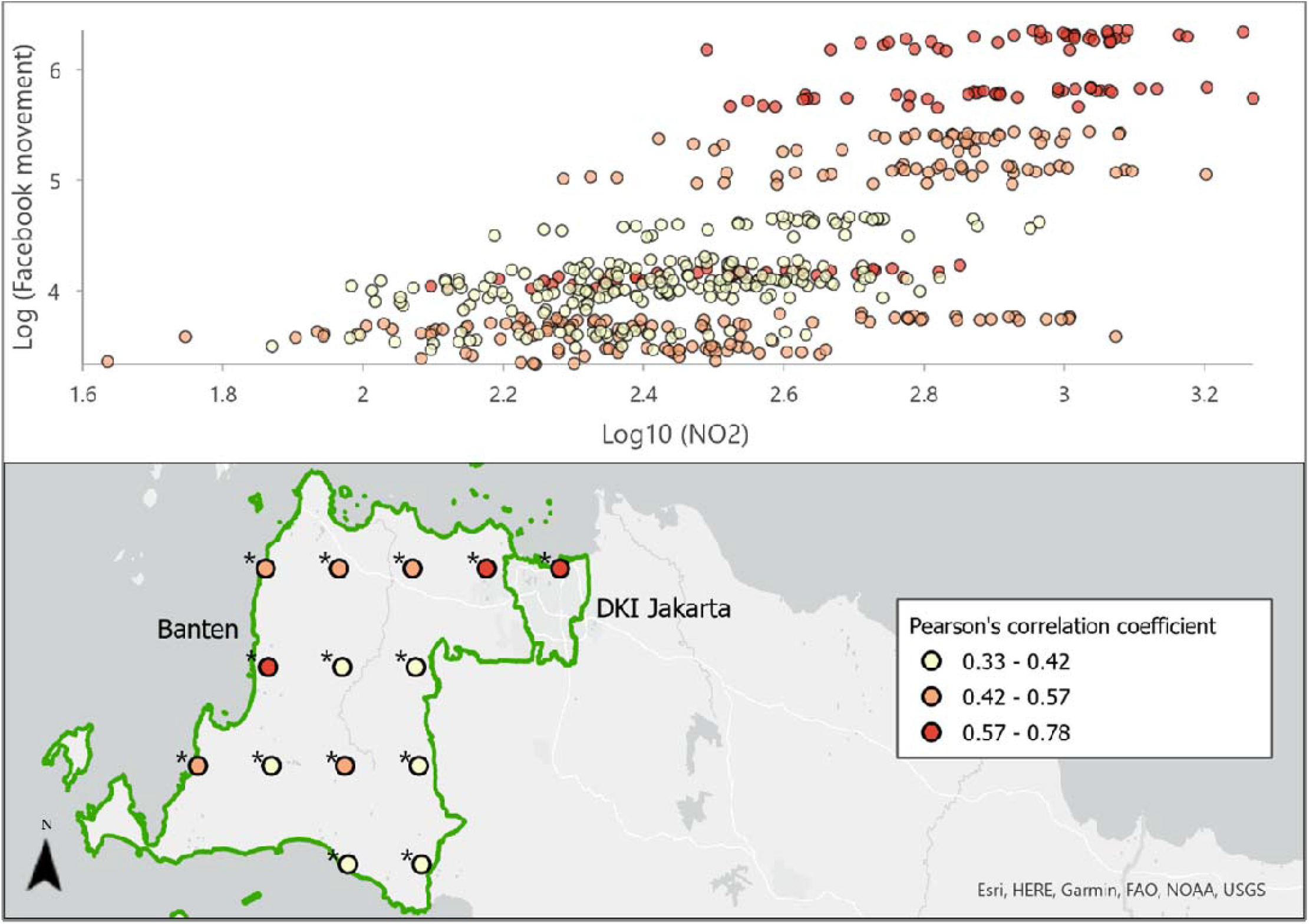
Correlation between Log(Facebook movement) and Log(NO_2_) in diminishing hot spots Indonesia.

**Figure 14.**
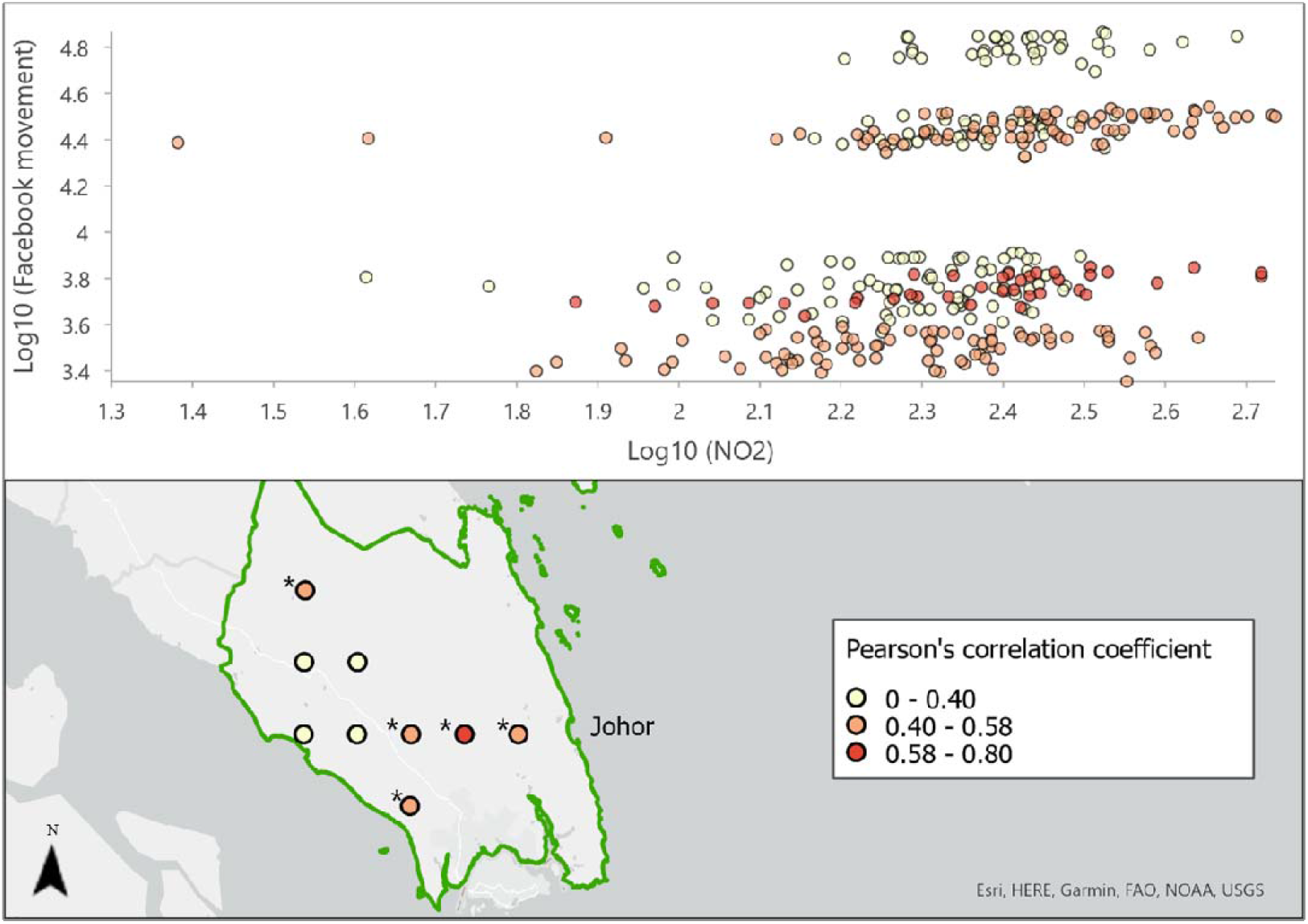
Correlation between Log(Facebook movement) and Log(NO_2_) in diminishing hot spots in Malaysia.

Overall, these spatially explicit exploration of the correlation between NO_2_ levels and mobility spatio-temporal trends revealed inconsistent patterns between June 2021 and February 2022, which approximates the period of the COVID-19 Delta and emerging Omicron variants. Further inspection of the correlation coefficients revealed that the observed correlations were only statistically significant for NO_2_ diminishing hot spots and the negatively correlated NO_2_ persistent and new hot spots. Therefore, we postulate that our SEA-wide EHSA results reinforce the findings from Li *et al*. (2022) where a national-scale spatial variability in correlations between NO_2_ levels and mobility was observed. This strongly indicates that mobility alone could not fully explain the resultant NO_2_ level changes over time and space.

### 3.2 NO_2_ prediction and its sensitivity to different influential factors

The previous two sections suggest that mobility is not the single influencing factor on the NO_2_ variation over time. We further explored the association between NO_2_ and other potential factors using the MLP model. Firstly, the correlation coefficients between each pair of variables were calculated and plotted as a heatmap (Figure 15). Log(facebook_movement) is the variable that has the strongest correlation with both Log(NO_2_) and NO_2_ (i.e., the correlation coefficient of 0.45 and 0.42, respectively). In comparison, Apple driving and walking trends have a much weaker correlation with Log(NO_2_) (i.e., the correlation coefficient of -0.13 and -0.02, respectively). However, Apple driving trend is strongly correlated to Apple walking trend, with a correlation coefficient of 0.73. This shows that Apple driving and Apple walking trends are very likely to increase or decrease together. Apart from the three mobility factors, longitude, latitude and temperature also showed a relatively strong correlation with NO_2_ and Log(NO_2_).

**Figure 15.**
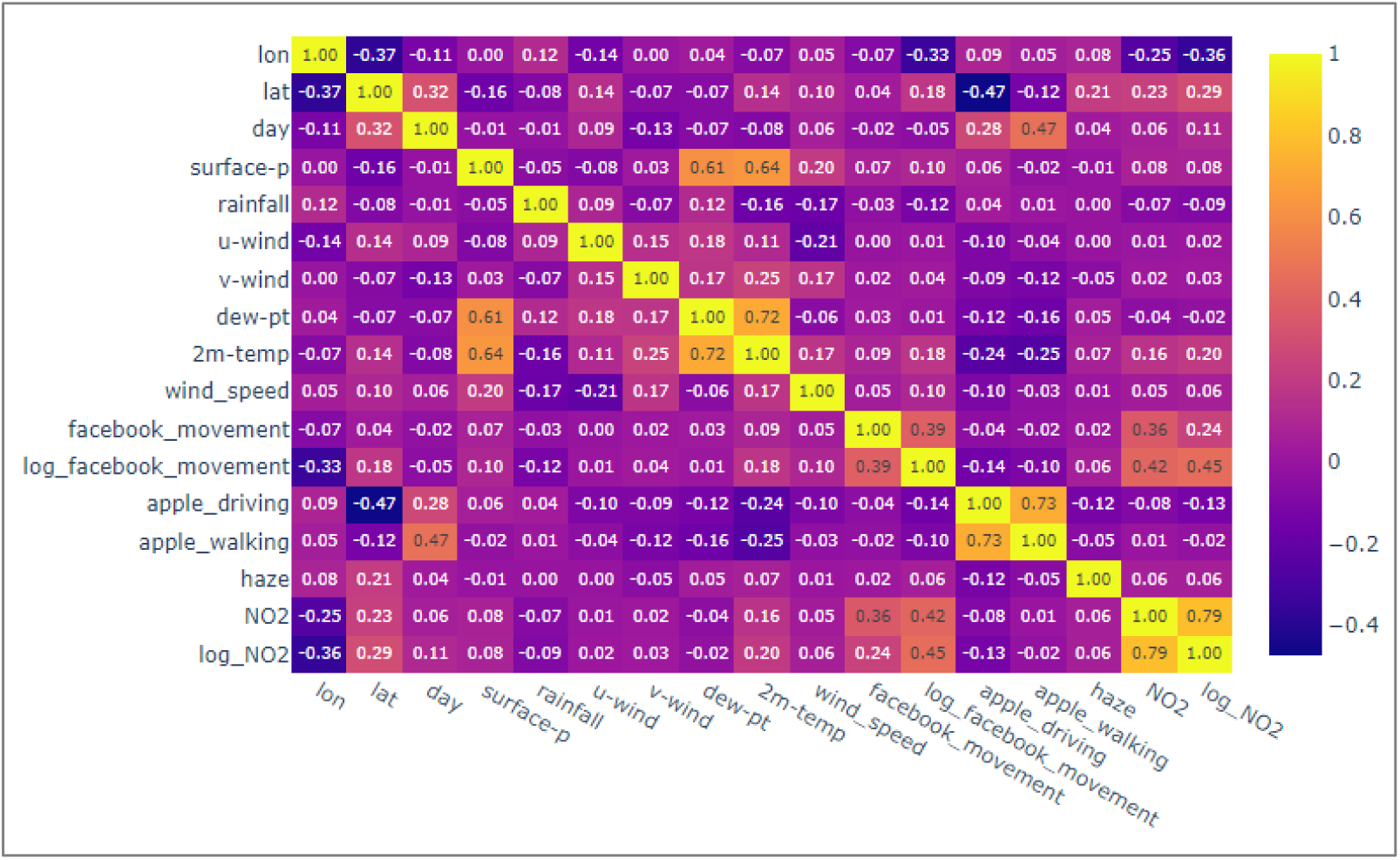
Correlation matrix between any pair of variables.

After calculating the coefficients, 15 features (Table 2) were used to predict the output NO_2_ in the MLP model. The model architecture consists of 18 hidden layers with a range of 8 to 64 nodes in each layer, and a total of 14,793 trainable parameters. Figure 16 plots the predicted and the measured ground-truth output values in both the training and testing datasets. A higher difference was observed at low Log(NO_2_) levels. Table 3 summarized the performance of both the training and testing datasets where the high similarity between their respective RMSE and MAE values suggests that no overfitting was found.

**Figure 16.**
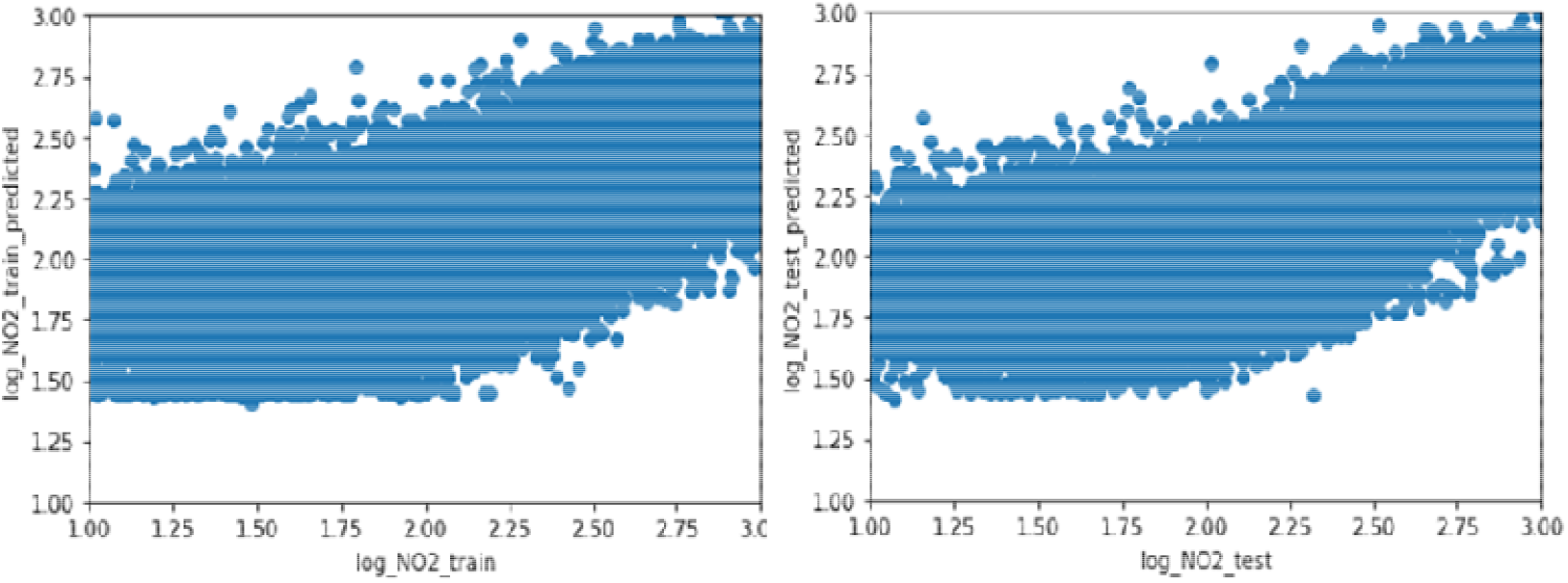
The predicted and measured Log(NO_2_) in the training (left) and testing (right) dataset.

**Table 3.** Performance of the MLP model on training and testing dataset.

|  | **RMSE**<br>**(NO<sub>2</sub>;**<br>**mol/m<sup>2</sup>)** | **RMSE**<br>**(Log(NO<sub>2</sub>))** | **MAE**<br>**(NO<sub>2</sub>;**<br>**mol/m<sup>2</sup>)** | **MAE**<br>**(Log(NO<sub>2</sub>))** | **R<sup>2</sup>**<br>**(NO<sub>2</sub>)** | **R<sup>2</sup>**<br>**(Log(NO<sub>2</sub>))** |
| --- | --- | --- | --- | --- | --- | --- |
| Train | 1.03e-05 | 0.22 | 5.03e-06 | 0.16 | 0.49 | 0.52 |
| Test | 1.02e-05 | 0.22 | 5.02e-06 | 0.16 | 0.49 | 0.52 |

The prediction error was evaluated by calculating the difference between the predicted and measured ground truth values in the testing dataset. The histogram of prediction error shows that most errors fall between -0.5 and 0.5 (Figure 17). A coloured distinction was used to differentiate observations with large prediction errors (> 0.5 or < -0.5) from those with small prediction errors (Figure 18). This distinction revealed that large errors concentrate at column densities lower than 1e-5.5 mol/m^2^ (or log(NO_2_) < 1.5). A measured NO_2_ column density of 1e-5.5 mol/m^2^ could be overestimated as 1e-5 mol/m^2^. On the other hand, measurements also have uncertainty. By comparing with ground measurements, the uncertainty in measurement was about 1e-5 mol/m^2^ (Tonion & Pirotti, 2022). While our prediction error occurred mostly at small NO_2_ column densities and was within the range of measurement uncertainty, this implies that the trained model performs well in predicting most of the NO_2_ levels and can generalise such prediction without overfitting the models.

**Figure 17.**
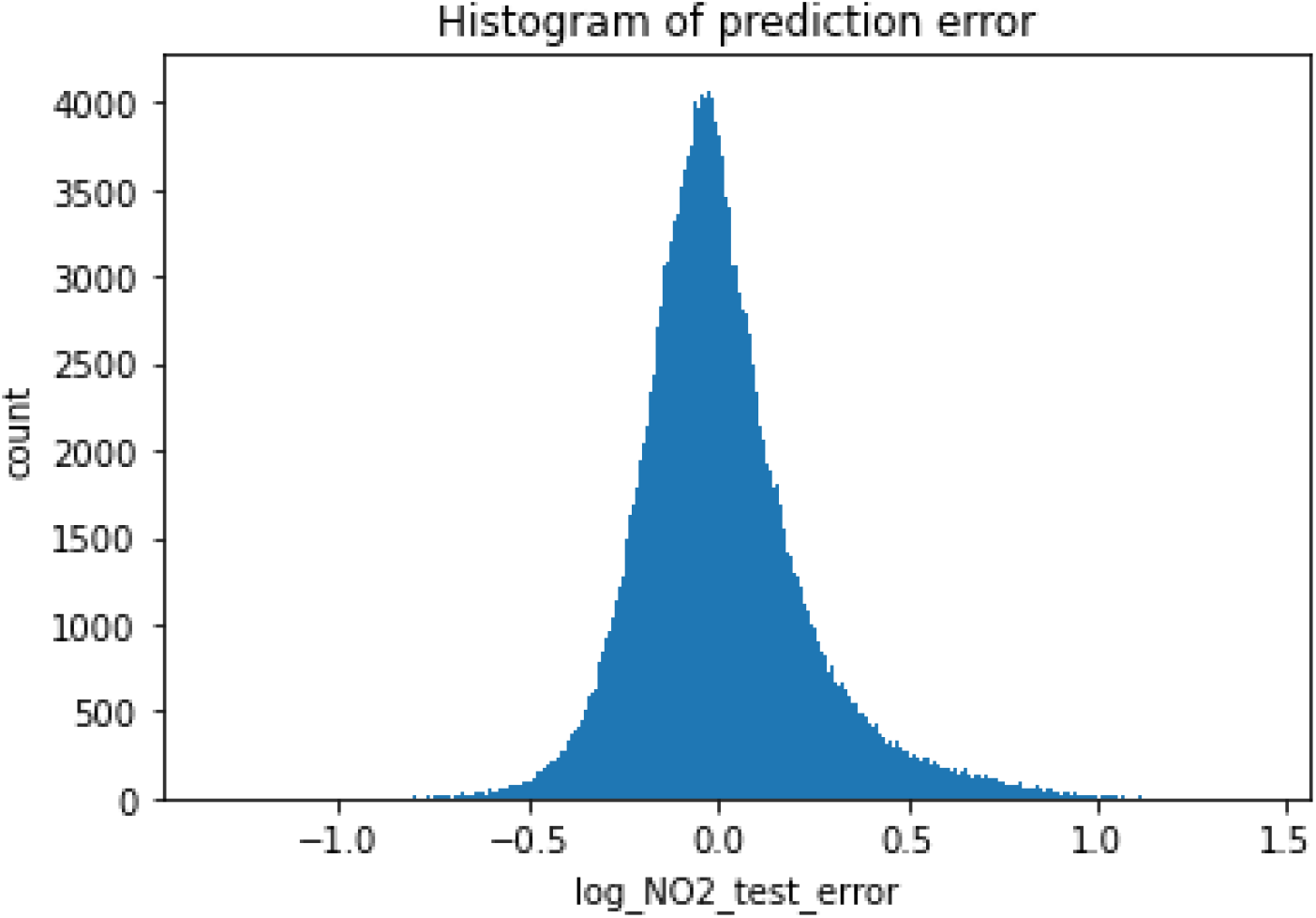
Histogram of prediction errors in the testing dataset.

**Figure 18.**
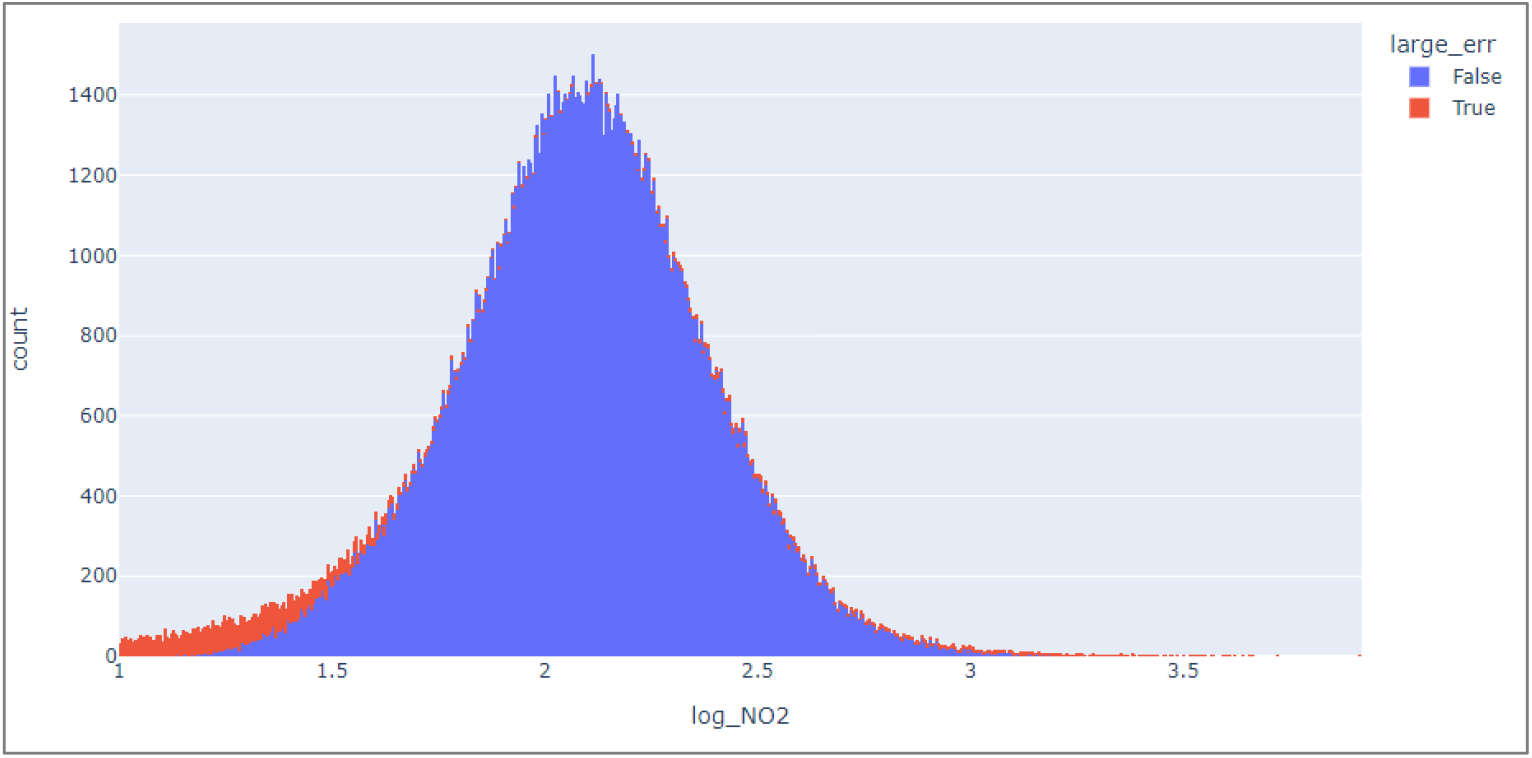
The distribution of large errors in the measured Log(NO_2_) in the testing dataset.

Subsequently, data records with high Log(NO_2_) were sliced as a new dataset to investigate its association with different parameters. This included 6362 Log(NO_2_) records ranging between 2.48 to 2.50 with small prediction error, representing a subset of data reliably predicted by the model. SHAP values were calculated for each parameter in each record representing the magnitude of impact each parameter has on the output Log(NO_2_).

The spatial (i.e., lat and lon) and temporal information (i.e., day) had the highest impacts on the output Log(NO_2_) according to the global interpretation summary (Figure 19). This reinforces the importance of spatial and temporal information in predicting NO_2_ levels. Besides spatial and temporal parameters, the next highest contributing parameters were mobility factors (i.e., apple_walking, apple_driving and facebook_movement). Specifically, the Apple moving trends had higher impacts on NO_2_ as compared to the Facebook movement. As mentioned, Facebook movement data has higher spatial resolution than Apple’s. Although the latter is coarser in resolution, it distinguished different travel modes (i.e., driving and walking). The comparatively higher impacts of Apple moving trends on Log(NO_2_) suggest that distinguishing different travel modes is important in predicting NO_2_. Additionally, other factors especially temperature also showed impacts to a certain extent on this model, although their impact is overall lower compared to spatio-temporal factors and most of the mobility factors.

**Figure 19.**
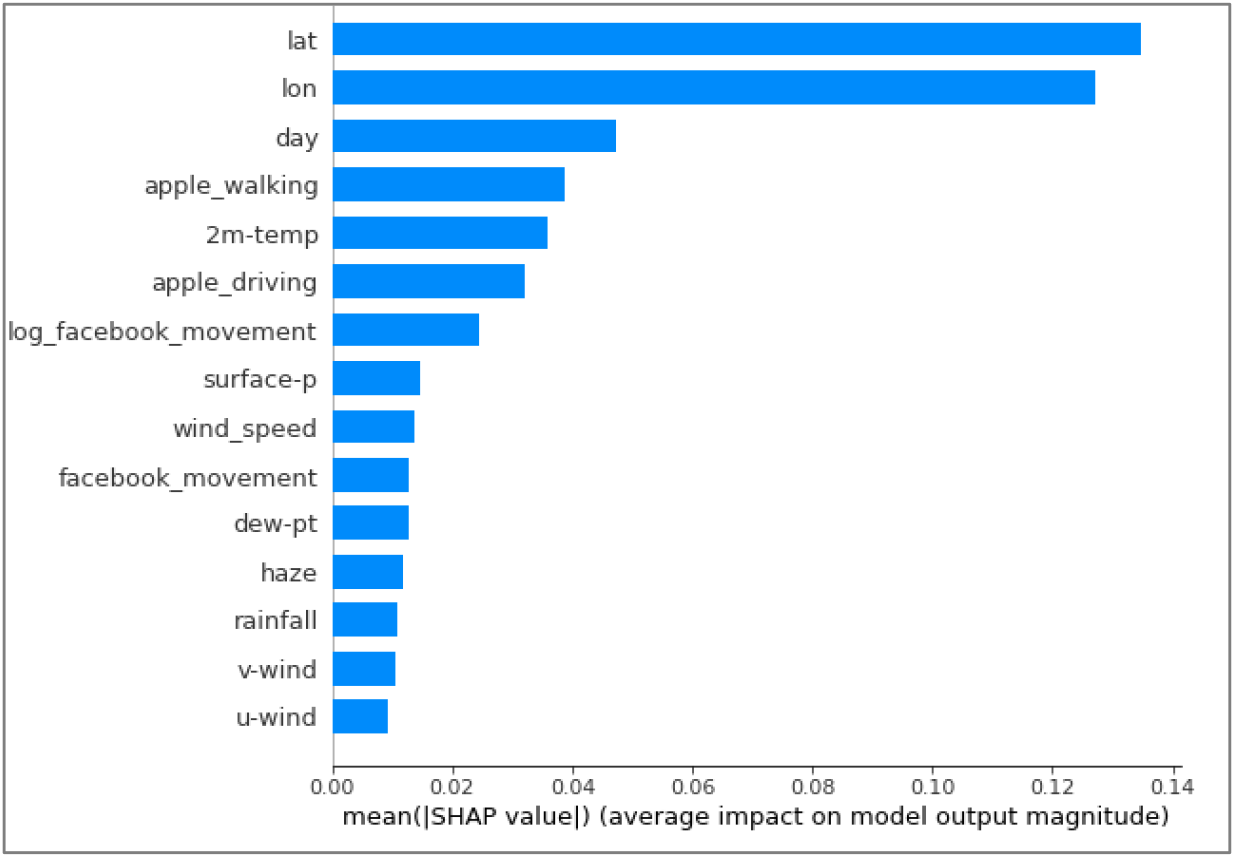
A summary plot of impacts of each parameter on Log(NO_2_).

The dependence plot in Figure 20 shows that when Log(facebook_movement) is below the average value (normalised to 0), both facebook movement and apple moving trend hardly have any impact on NO_2_. In other words, the impact of change in mobility parameters on NO_2_ is more detectable at higher movement level. On the other hand, this impact on NO_2_ could be different at the same total movement level. For instance, when Log(facebook_movement) was above the mean value, the blue points (lower level of Apple driving trend) have lower SHAP values compared to the pink points (higher level of Apple driving trend). This suggests that when total mobility is contributed less by the driving trend, an increase in total mobility could be contributed more by the walking trend instead. Therefore, there tends to be a lower resultant impact on NO_2_ levels.

**Figure 20.**
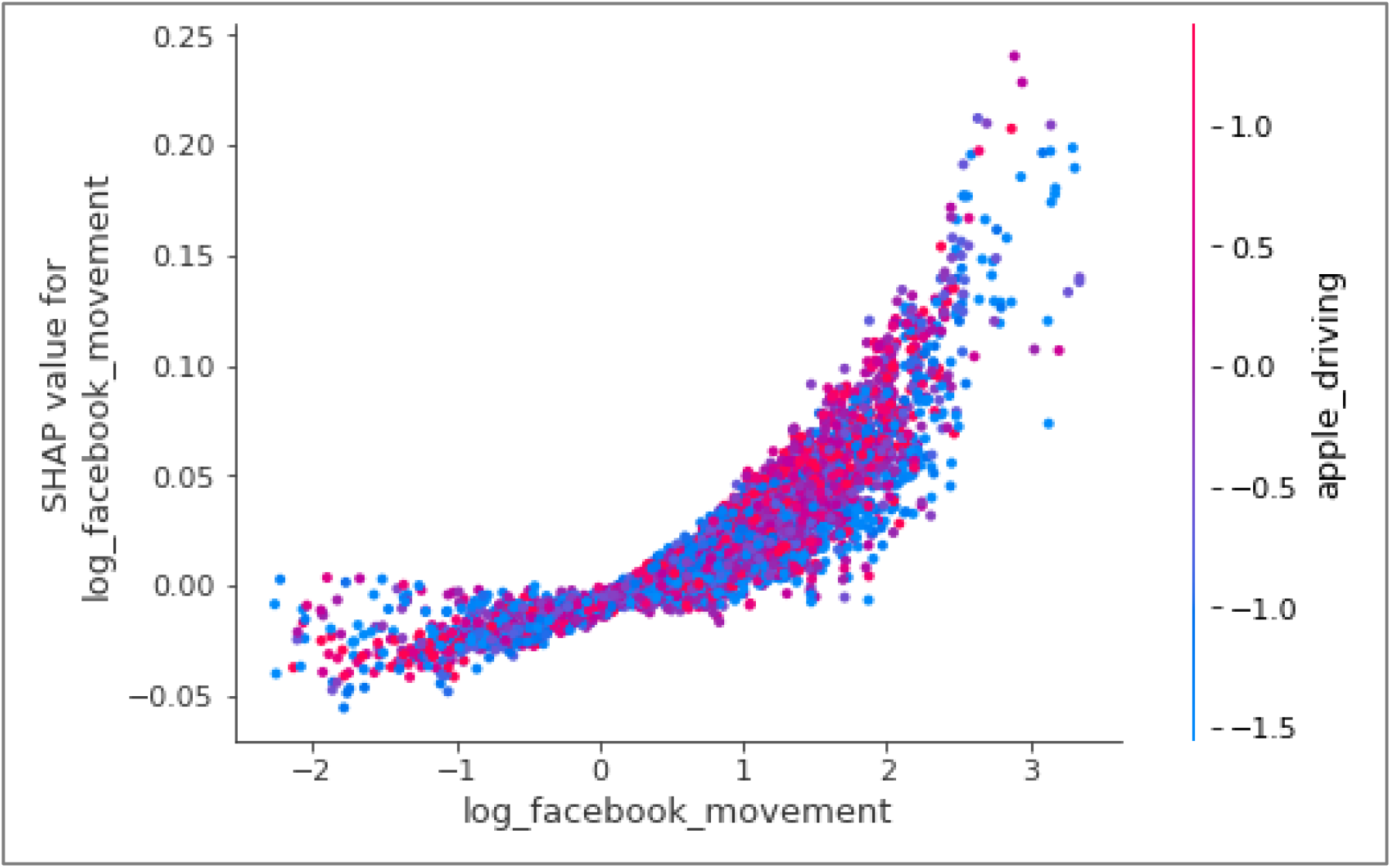
Impact of Log(facebook_movement) on Log(NO_2_) represented by the SHAP values at different Log(facebook_movement) values along the x-axis, and its interactions with Apple driving trend (in colour).

In addition, the impact of Apple walking trend on Log(NO_2_) in Figure 21 shows that there is a negative impact on Log(NO_2_) when the walking trend decreases. The different interactions of Apple driving and Apple walking indicated that this negative impact on Log(NO_2_) from a decreased walking trend could be attributed to the concurrent decrease in Apple driving trend (in blue colour).

**Figure 21.**
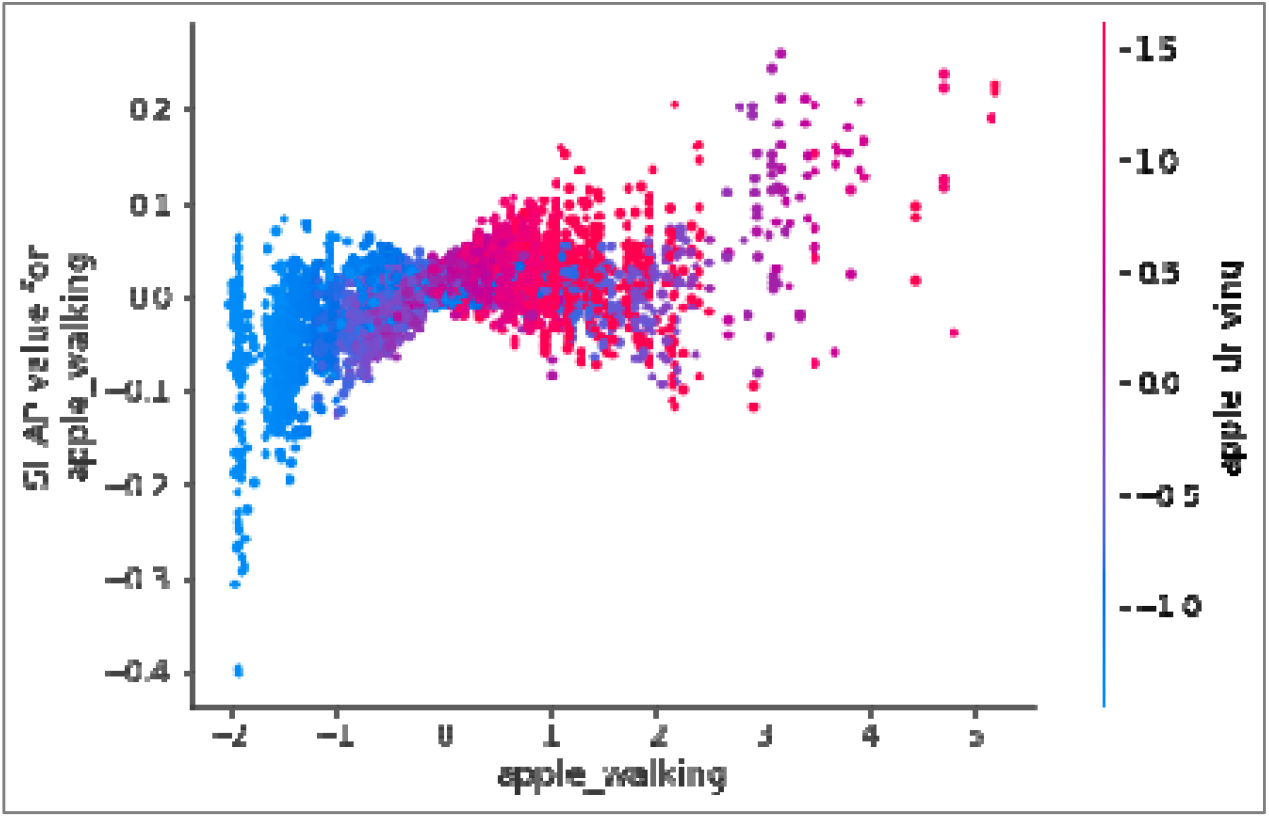
Impact of Apple walking trend on Log(NO_2_) represented by the SHAP values at different apple_walking values along the x-axis, and its interactions with Apple driving trend (in colour).

## 4 Conclusions

The various lockdowns and movement restrictions during the COVID-19 pandemic present an opportunity to study how air pollution varies along with the decrease of mobility. Although existing studies have conducted analysis during the lockdowns in early 2020 and observed a positive correlation between NO_2_ and mobility. Given that their study period was confined to a short time period, their observation may be subject to other under-examined factors. Therefore, we investigated the variability of NO_2_ at a very large spatial scale with diverse human movement patterns in a two-year study time period. We found that mobility showed decreasing trend during the COVID-19 pandemic in Indonesia. However, the decreased mobility did not generally result in concurrent decrease in NO_2_ during the two years. Our EHSA results revealed the spatio-temporal heterogeneity in NO_2_ levels and mobility in SEA during June 2021 and February 2022. The variability in Pearson’s correlation coefficient further indicated the absence of a definitive correlation between NO_2_ and Facebook movement. The subsequent MLP result suggests that mobility with distinguished travel modes substantially influenced the MLP model and NO_2_ prediction. Moreover, the impact of mobility on NO_2_ varies with mobility level. When mobility is below the average level, its impact on NO_2_ becomes negligible. This may be another reason why the decrease in mobility did not result in any expected correlated decrease in NO_2_ in general.

Our findings denoted that the reduced mobility during COVID-19 pandemic did contribute to the variation of NO_2_. Nevertheless, other meteorological factors especially temperature also had considerable impacts and should be considered along with mobility when attempting to understand NO_2_ variation. Although SEA countries are gradually loosening movement and other restrictions to restore industry activities and economy (Luo *et al*., 2022), traffic control is still a recommended solution for regions suffering from poor air quality. Additionally, our model provides an accurate NO_2_ prediction method, which could be adopted as a health risk warning system in preventing possible exposure to high concentration of NO_2_.

There are three main limitations in this study and further investigations are recommended. Firstly, our study mainly estimated the movement value in adjacent study grids, excluding long distance travels. Although daily commuting in adjacent grids accounts for most of the movement value (93.33 percent), this would cause some uncertainty in actual movement volume estimation. Future studies could adopt other datasets that include specific mobility trajectories to account for long distance travel patterns and reduce uncertainty. Secondly, as this study seeks to examine the spatio-temporal changes in NO_2_ levels, only specific patterns of hot spots (i.e., new, intensifying, persistent and diminishing hot spot) were selected for correlation analyses. Other studies could examine the correlation between NO_2_ and mobility levels for other patterns of hot spots and even the cold spots to potentially elicit more insights. Finally, this study did not adopt a spatio-temporal machine learning model due to the discontinuous Facebook movement data (i.e., most countries have missing data before May 2021) during the two-year study period. Future studies are recommended to adopt a hybrid model (e.g., convolutional neural network (CNN) and long short-term memory (LSTM)) for NO_2_ prediction. Nonetheless, it presented timely and novel evidence that the spatial and temporal information were strong factors in explaining NO_2_ levels. Future scholars are recommended to adopt a hybrid model (e.g., convolutional neural network (CNN) and long short-term memory (LSTM)) in NO_2_ prediction to account for the interactions of NO_2_ with time and space.

Source codes related to this study can be found on a public GitHub repository (https://github.com/liyangyang515/Spatio-Temporal-Patterns-of-NO_2_-and-Mobility-Through-the-Variants-of-COVID-19-in-SEA).

## Data Availability

All data produced in the present study are available upon reasonable request to the authors

https://partners.facebook.com/

https://earthengine.google.com/

https://trends.google.com/trends/

## Acknowledgements

The Facebook Movement between Tiles data are not publicly available, the permission of usage was granted by Facebook Data for Good Partner Program.

## Appendix

**Appendix A:**
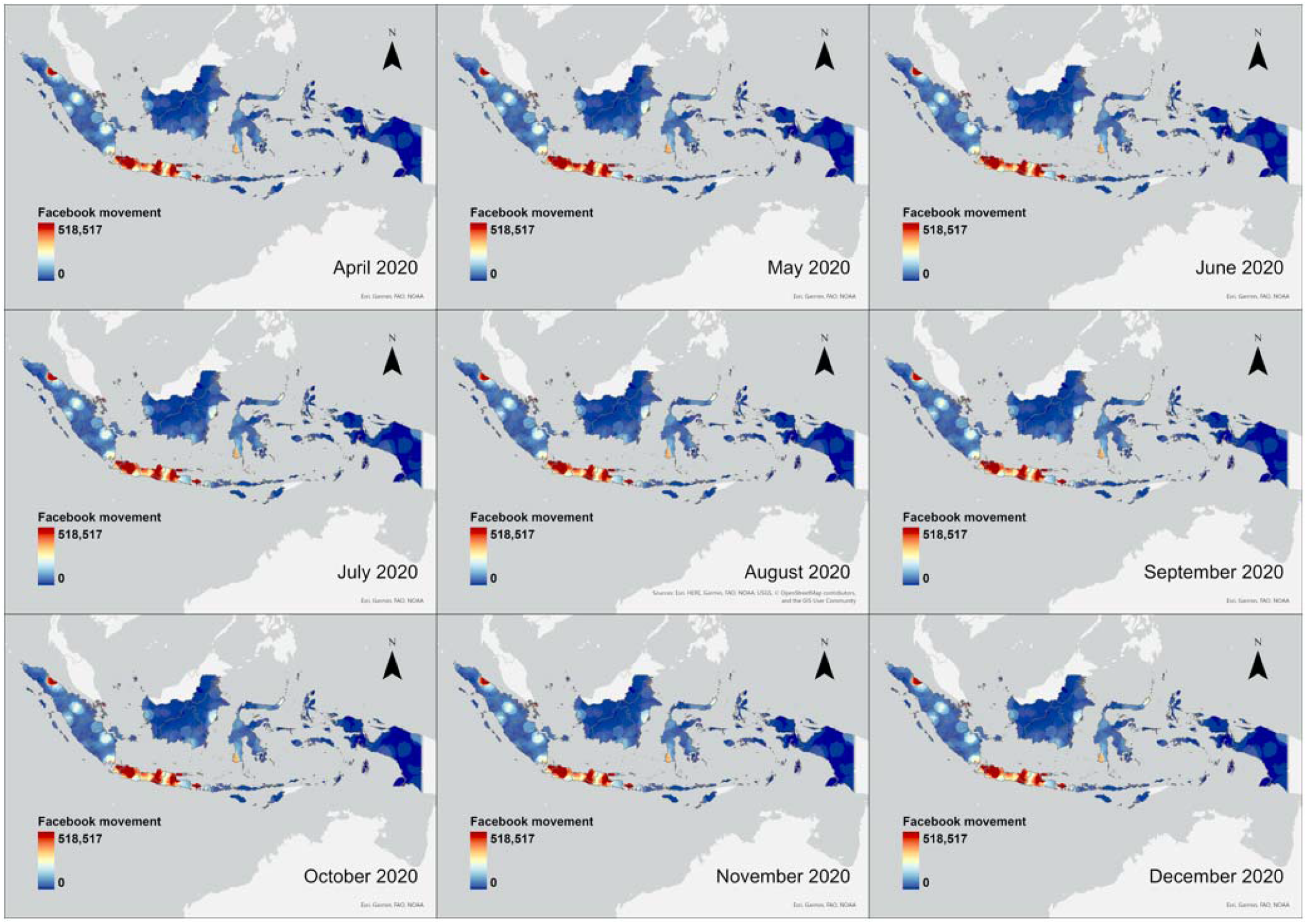
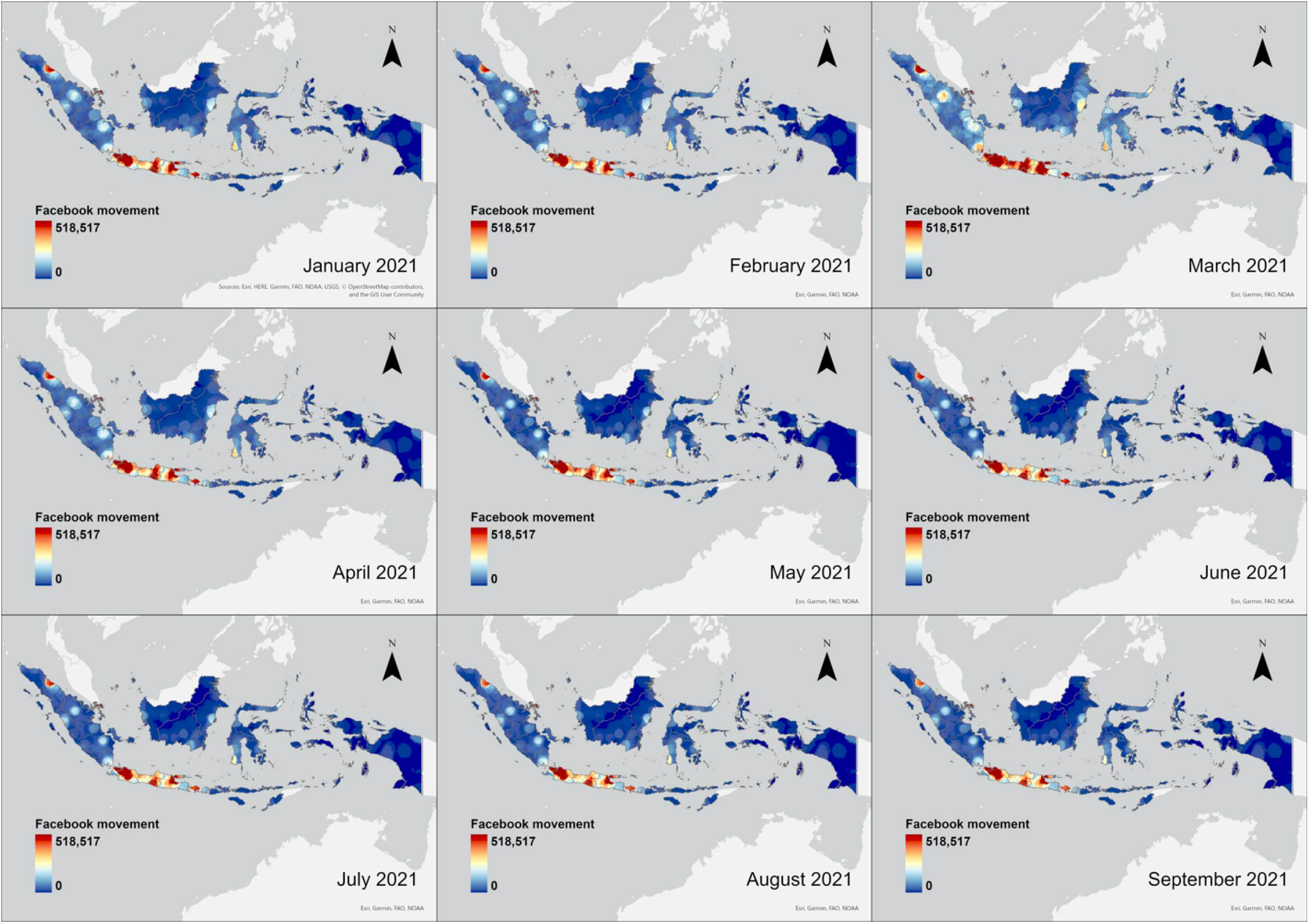
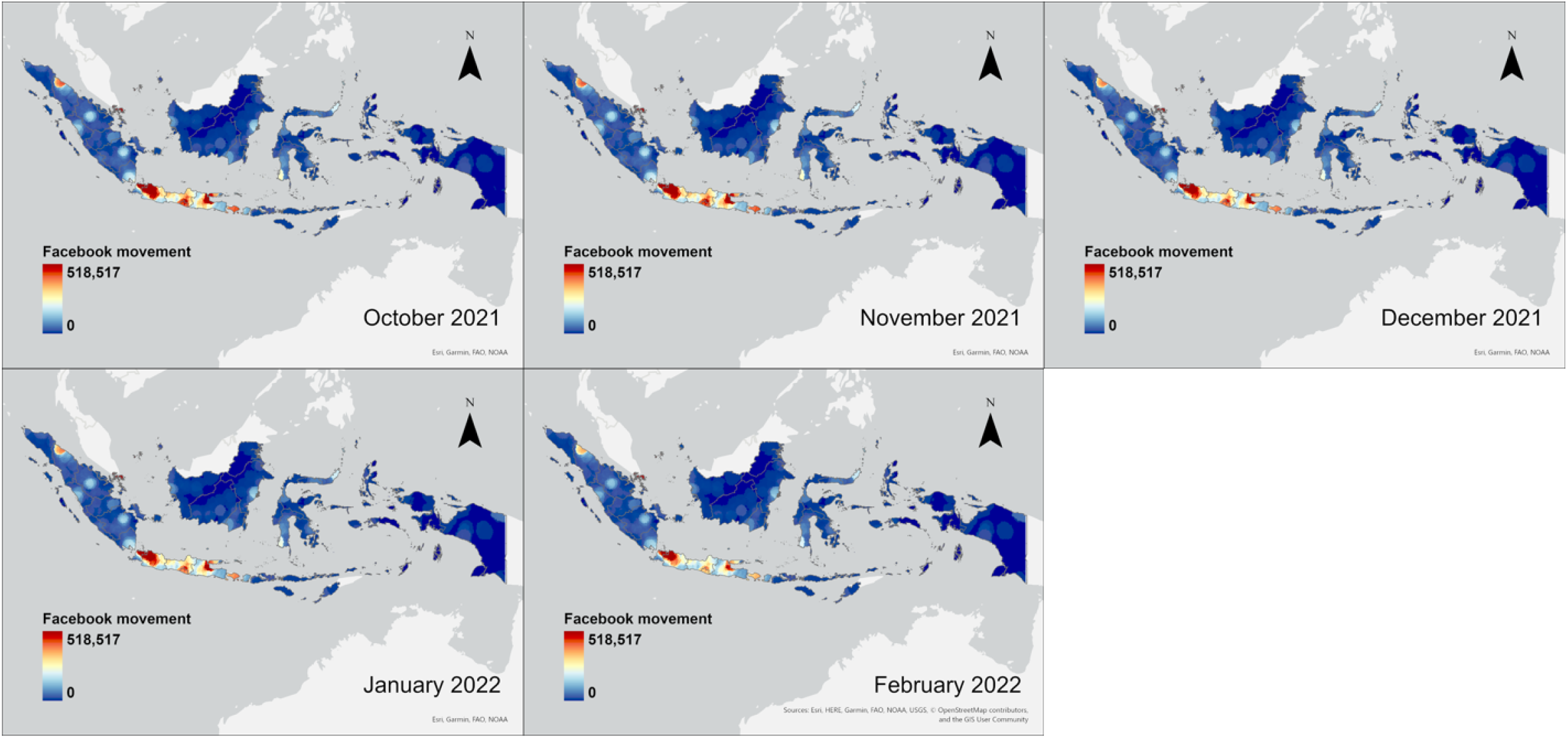
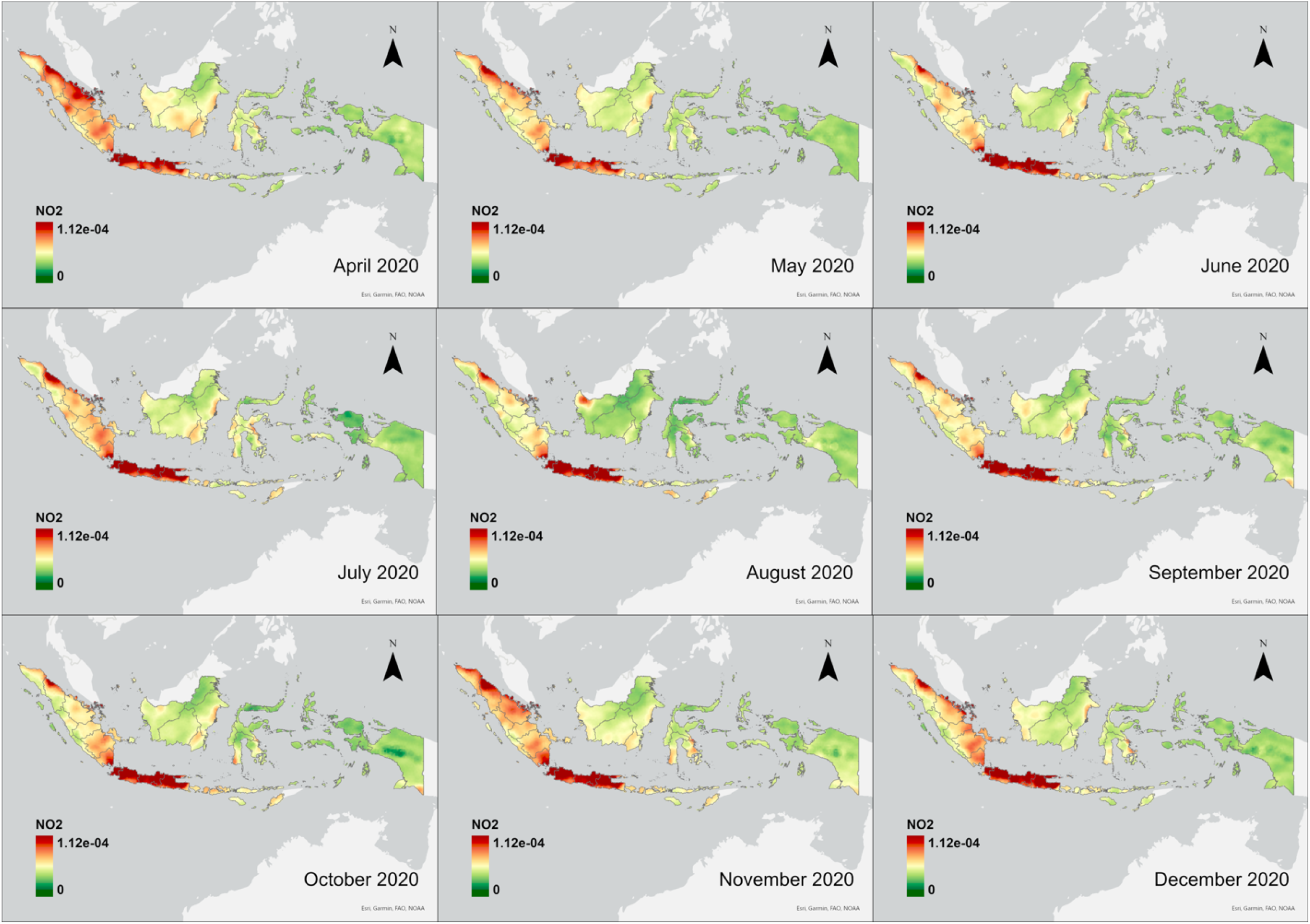
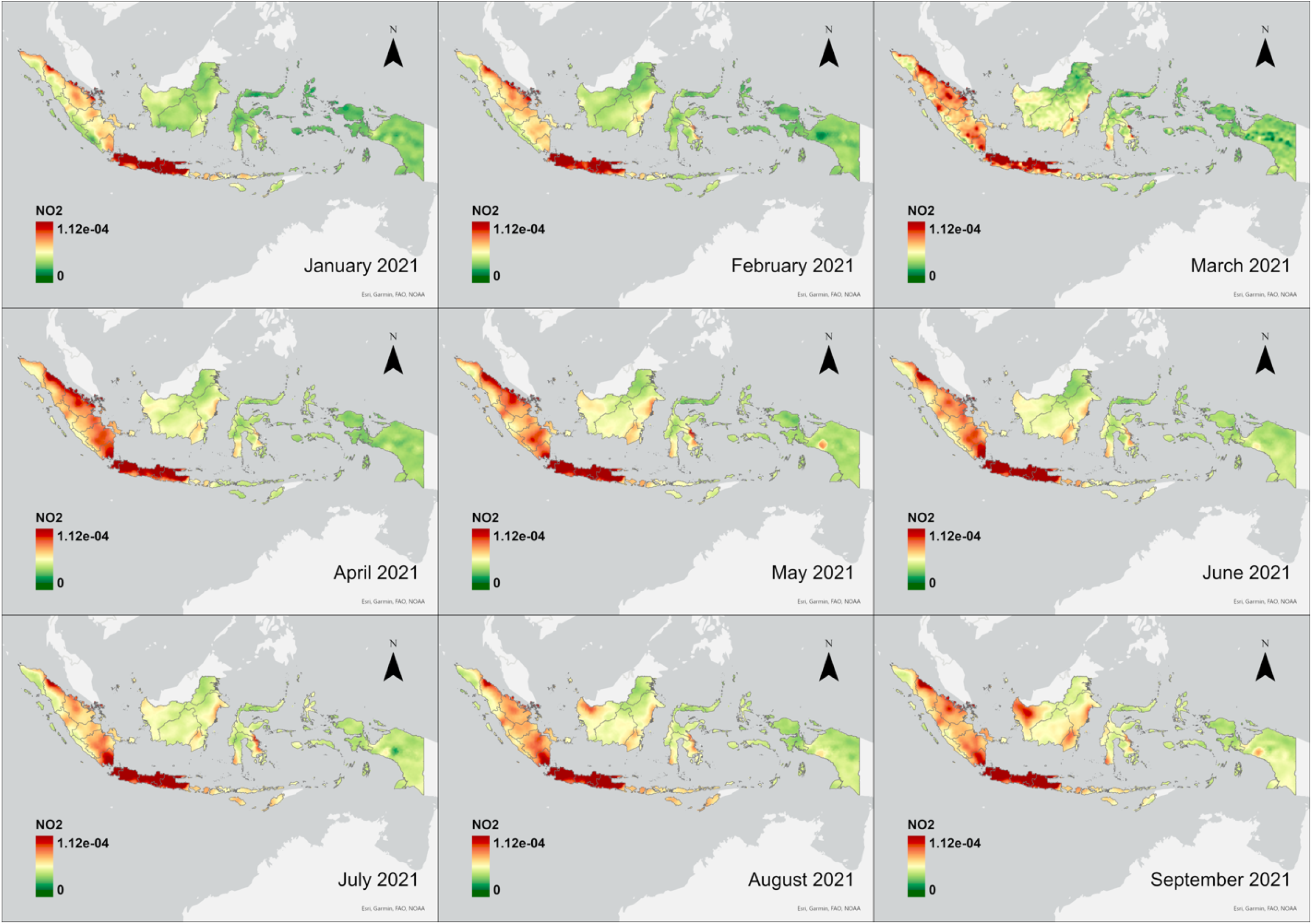
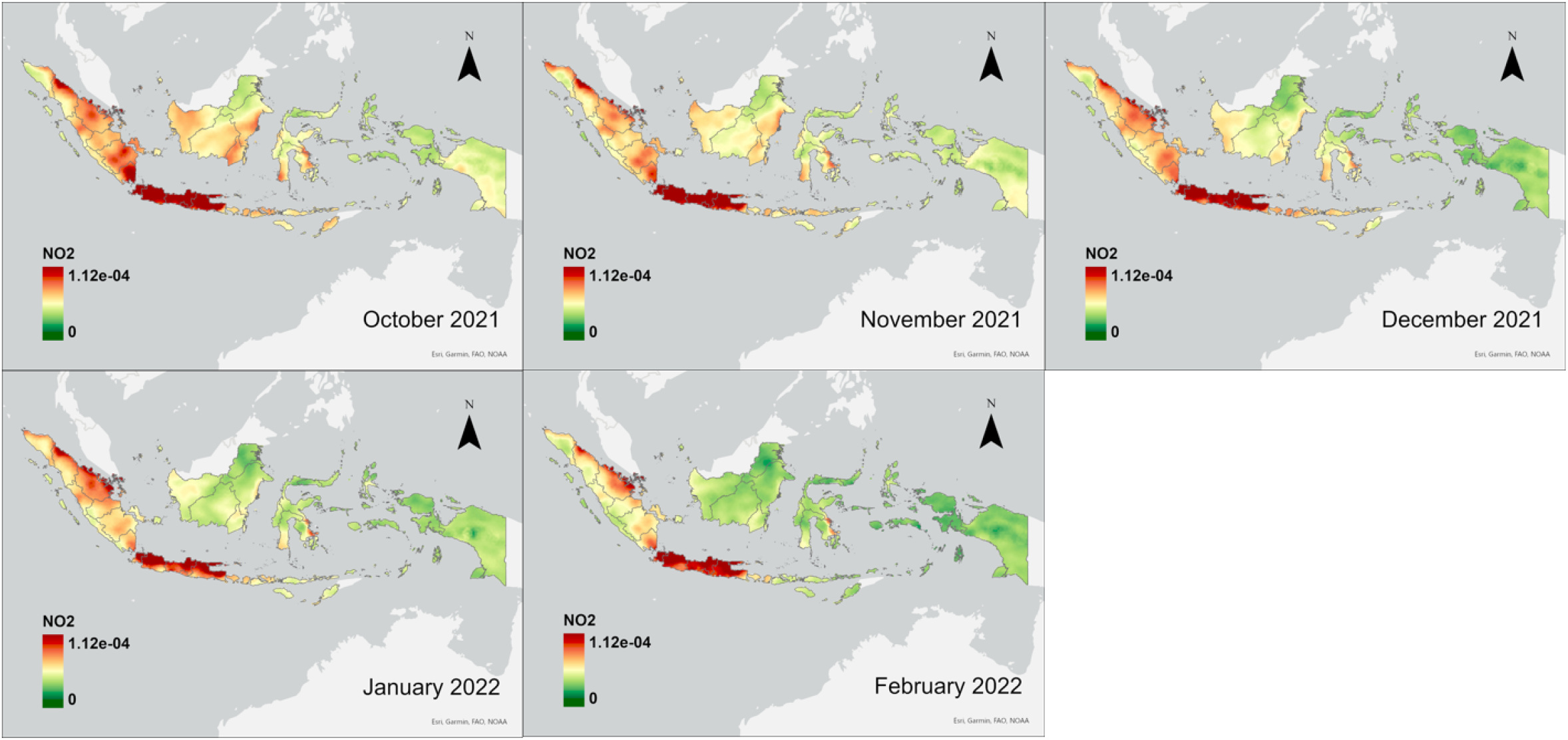
Distribution and Temporal Variation of Facebook movement and NO_2_ in Indonesia, between Apr 2020 and Feb 2022.

## Notes

### Competing Interest Statement

The authors have declared no competing interest.

### Funding Statement

This study did not receive any funding

